# Investigating the genetic relationship between depression symptoms and Alzheimer’s Disease in clinically diagnosed and proxy cases

**DOI:** 10.1101/2023.06.05.23290588

**Authors:** Lachlan Gilchrist, Thomas P. Spargo, Rebecca E. Green, Jonathan R.I. Coleman, David M. Howard, Jackson G. Thorp, Brett Adey, Jodie Lord, Helena L. Davies, Jessica Mundy, Abigail ter Kuile, Molly R. Davies, Christopher Hübel, Shannon Bristow, Sang Hyuck Lee, Henry Rogers, Charles Curtis, Gursharan Kalsi, Ryan Arathimos, Anne Corbett, Clive Ballard, Helen Brooker, Byron Creese, Dag Aarsland, Adam Hampshire, Latha Velayudhan, Thalia C. Eley, Gerome Breen, Alfredo Iacoangeli, Sulev Koks, Cathryn M. Lewis, Petroula Proitsi, Alzheimer’s Disease Neuroimaging Initiative AddNeuroMed GERAD1 Consortium and GERAD1 Consortium

## Abstract

Depression is a risk factor for Alzheimer’s disease (AD), but evidence for their genetic relationship is mixed. Assessing depression symptom specific genetic associations may better clarify this relationship.

Using data from the UK Biobank, the GLAD Study and PROTECT, we performed the largest genome-wide meta-analyses (GWAS) of the nine depression symptom items, plus their sum score, on the Patient Health Questionnaire (PHQ-9) (GWAS equivalent N: 224,535—308,421). We assessed global/local genetic correlations and statistical colocalisation between depression phenotypes and AD across six AD GWAS with varying proportions of clinical and proxy (family history) case ascertainment. We assessed bi-directional causal associations using Mendelian randomisation (MR) and the predictiveness of depression phenotype polygenic risk scores (PRS) for AD case/control status in three clinical AD cohorts.

Our GWAS meta-analyses identified 37 genomic risk loci across the ten depression symptom phenotypes. Of the 72 global genetic correlation tests conducted between depression/depression symptoms and AD, 20 were significant at p^FDR^≤ 0.05. Only one significant genetic correlation was identified with AD GWAS containing clinical-only cases. Colocalisation was not identified at loci contains local genetic correlation but was identified in the region of transmembrane protein 106B (TMEM106B) between multiple depression phenotypes and both clinical-only and clinical+proxy AD. MR and PRS analyses did not yield statistically significant results.

Our findings do not demonstrate a causal role of depression/depression symptoms on AD and suggest that previous evidence of their genetic overlap may be driven by the inclusion of proxy cases/controls. However, the identification of colocalisation at TMEM106B warrants further investigation.

## 1. INTRODUCTION

Epidemiological studies suggest that a diagnosis of depression is a risk factor for the later development of dementia^1–6^, of which Alzheimer’s disease (AD) is the most common form, accounting for approximately 80% of the over 40 million global cases^7^. Establishing the underlying mechanisms by which depression confers increased risk for AD offers a pathway by which new interventions might be implemented and the global dementia burden reduced^8^.

As twin studies have demonstrated, both depression and AD are substantially heritable – approximately 40% and 80% respectively^9, 10^. Further, large-scale genome-wide association studies (GWAS) have demonstrated high polygenicity, identifying over 70 genomic risk loci for AD and nearly 200 for depression^11–17^. It is therefore possible that their phenotypic association is in part due to shared genetic architecture. However, results from previous investigations into the genetic overlap between the two disorders have been mixed. For example, some findings indicate non-significant genetic overlap^18, 19^, others a significant – if modest – genetic correlation of ∼16-17% and a risk increasing causal effect of depression on AD^20–22^.

According to the Diagnostic and Statistical Manual of Mental Disorders (DSM-5), diagnosis of a major depressive episode requires the presence of at least five of a possible nine symptoms for ≥ 2 weeks, including one of the two cardinal symptoms – depressed mood or anhedonia^23^. Potentially hundreds of symptom combinations are possible to meet this diagnosis criteria^24–26^. As such, this heterogeneity poses challenges to researchers seeking to better understand differences in the genetic contribution to depression and its subtypes^27, 28^. However, the decomposition of depression into individual symptoms has provided insight into unique patterns of genome-wide significant loci and cross-trait genetic associations, as demonstrated in a recent GWAS of depression symptoms on the Patient Health Questionnaire (PHQ-9) by Thorp et al.^29^.

A number of studies suggest that anhedonia may be a better predictor of dementia than depressed mood^30–32^. Further, several depression symptoms, such as appetite changes, psychomotor dysfunction, and sleep disruption are commonly observed in non-depressed dementia patients^33–35^. Taking this into account alongside the mixed nature of previous findings examining the genetic overlap between depression and AD, it is possible that leveraging depression symptom level genetic information may offer greater insight into the disorders shared genetic architecture.

However, any association between depression and AD must also consider the potential influence of differences in case/control ascertainment in AD GWAS. A review by Escott-Price et al.^36^ notes that recent large-scale AD GWAS contain a relatively small proportion of clinically ascertained cases/controls, with a large percentage of cases ascertained by proxy – that is, cases and controls are defined as individuals with and without a self-reported parental history of AD/dementia, respectively. The combination of clinical and proxy samples in AD GWAS meta-analyses has proved an effective way of boosting sample size and variant discovery^15–17, 37^. However, evidence suggests that this has come at the expense of specificity in regard to genomic risk loci and an apparent stagnation in the percentage of variance explained by common variants^36^. Most importantly for cross-trait analysis, recent studies indicate that the direction of Mendelian Randomisation (MR) causal estimates for AD risk factors on AD can be in the opposite direction depending on whether the AD outcome GWAS contains clinical and proxy cases/controls or is more strictly clinically ascertained^38–40^.

To address these points, we perform the first and largest genome-wide meta-analyses of PHQ-9 depression symptom items using data from the Genetic Links to Anxiety and Depression (GLAD) Study^41^, the PROTECT Study^42, 43^ and two questionnaires from UK Biobank (UKB)^44^. We obtained summary statistics from previous large-scale GWAS for clinical^11^ and broad^12^ depression, and six AD GWAS. Specifically, three with clinical+proxy case/control ascertainment^15–17^, one with proxy-only^37^, and two with clinical-only^14, 45^. We used these GWAS to assess the presence, strength and differences in genetic overlap between depression, depression symptoms and AD, with the additional aim of better understanding the influence of different AD case ascertainment strategies on associations.

## 2. METHODS

For an analysis flowchart, see Figure 1.

**Figure 1:**
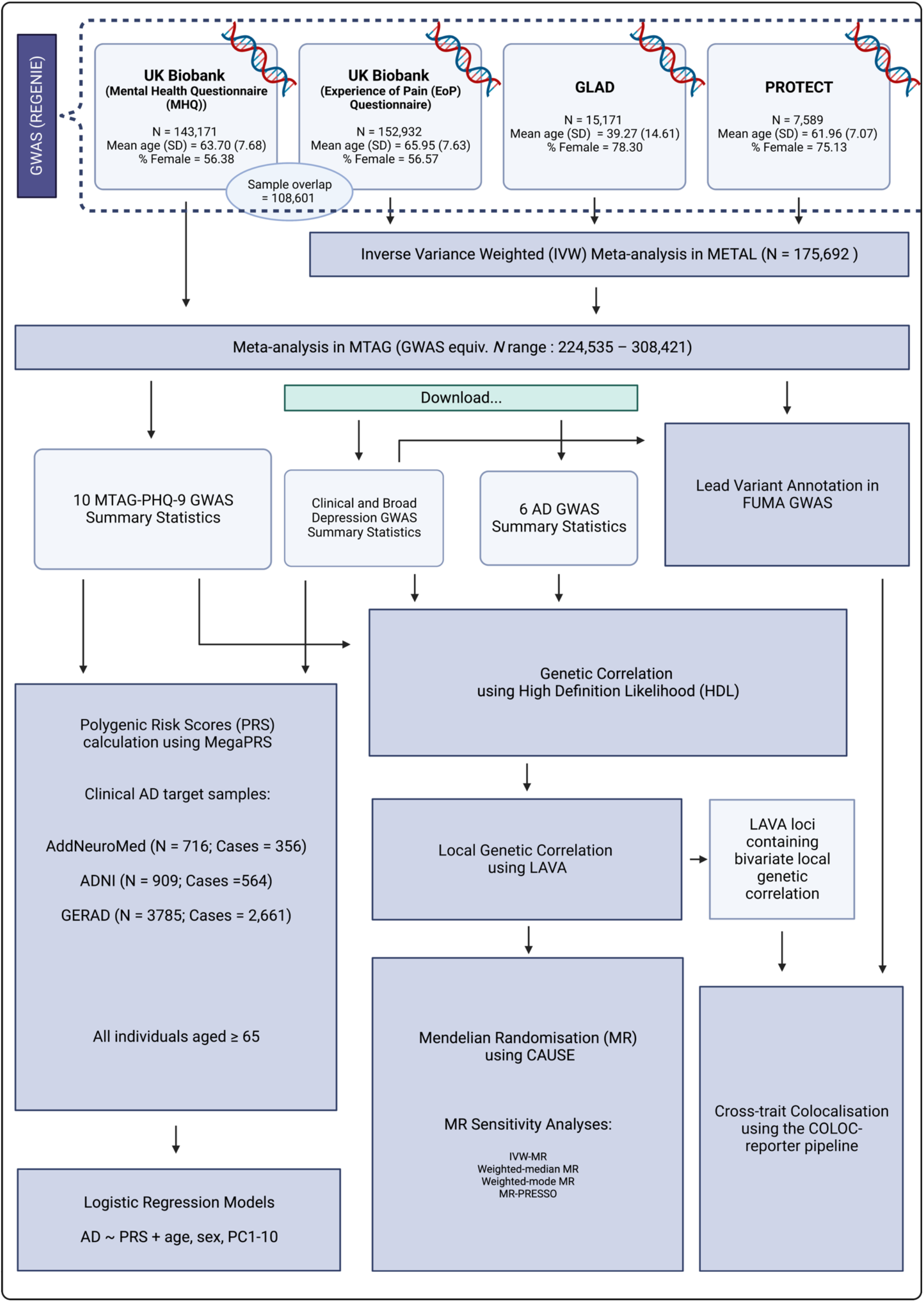
Analysis flowchart for the present study describing the analyses undertaken following genetic and phenotypic quality control (see: Methods) for each of the depression symptom items of the Patient Health Questionnaire (PHQ-9) in each of the four samples – UK Biobank (Mental Health Questionnaire and Experience of Pain Questionnaire), the Genetic Links in Anxiety and Depression (GLAD) Study and the PROTECT Study. GWAS = Genome-wide association analysis; AD = Alzheimer’s Disease; MTAG = Multi-trait Analysis of GWAS; FUMA = Functional Mapping and Annotation; LAVA = Local Analysis of [co]Variant Association; ADNI = Alzheimer’s Disease Neuroimaging Initiative; PC = genetic principal components

### 2.1 GWAS in the UK Biobank, GLAD and PROTECT

#### 2.1.1 Patient Health Questionnaire-9 (PHQ-9) phenotypes

The PHQ-9 is a well-validated clinical screening questionnaire used to assess depression symptom severity on nine individual symptoms in the DSM-IV^46, 47^. The severity of each symptom is measured by the self-reported persistence of that symptom over the preceding two weeks on a scale of 0 to 3. Scores of 3 indicate an individual experienced that symptom nearly every day, 2 indicates an individual experienced that symptom more than half the days, 1 indicates an individual experienced that symptom for several days and 0 indicates no experience of that symptom at all. The sum of an individual’s scores over all nine items (sum-score) ranges from 0-27. For an overview of the PHQ-9 items and response distribution for each sample, see Table 1. Sum-score distributions can be seen in Supplementary Table 1.

**Table 1:** Patient Health Questionnaire (PHQ-9) derived depression symptom phenotypes, their corresponding question on the PHQ-9, and the distribution and percentage of responses for each of the four samples on which genome-wide association analysis was performed.

| Depression Symptom | PHQ-9 Question: | UK Biobank Data Field (MHQ/EoP) | Cohort | PHQ-9 Response |  |  |  |
| --- | --- | --- | --- | --- | --- | --- | --- |
|  | *How often in the last 2 weeks have you been bothered by any of the following problems?* |  |  | *0 (Not at all)* | *1 (Several days)* | *2 (more than half the days)* | *3 (Nearly every day)* |
| Anhedonia | Little interest or pleasure in doing things | 20514/120104 | UK Biobank (MHQ) | 117045 (81.75%) | 21088 (14.73%) | 2965 (2.07%) | 2073 (1.45%) |
|  |  |  | UK Biobank (EoP) | 116330 (76.07%) | 29743 (19.45%) | 4115 (2.69%) | 2744 (1.79%) |
|  |  |  | GLAD | 4108 (27.08%) | 6429 (42.31%) | 2502 (16.49%) | 2142 (14.12%) |
|  |  |  | PROTECT | 6194 (81.62%) | 1130 (14.89%) | 127 (1.67%) | 138 (1.82%) |
|  |  |  | **Total** | **243677 (76.42%)** | **58380 (18.31%)** | **9709 (3.04%)** | **7097 (2.23%)** |
| Appetite Changes | Poor appetite or overeating | 20511/120108 | UK Biobank (MHQ) | 117505 (82.07%) | 18521 (12.94%) | 3860 (2.70%) | 3285 (2.29%) |
|  |  |  | UK Biobank (EoP) | 121071 (79.17%) | 22432 (14.67%) | 5404 (3.53%) | 4025 (2.63%) |
|  |  |  | GLAD | 4242 (27.96%) | 4459 (29.39%) | 3057 (20.15%) | 3413 (22.50%) |
|  |  |  | PROTECT | 6364 (83.86%) | 971 (12.79%) | 145 (1.91%) | 109 (1.44%) |
|  |  |  | **Total** | **249182 (78.15%)** | **46383 (14.55%)** | **12466 (3.90%)** | **10832 (3.40%)** |
| Concentration Problems | Trouble concentrating on things | 20508/120110 | UK Biobank (MHQ) | 117951 (82.07%) | 20386 (14.24%) | 2713 (1.89%) | 2121 (1.48%) |
|  |  |  | UK Biobank (EoP) | 123920 (81.03%) | 23441 (15.33%) | 3258 (2.13%) | 2313 (1.51%) |
|  |  |  | GLAD | 4665 (30.75%) | 5200 (34.28%) | 2891 (19.06%) | 2415 (15.92%) |
|  |  |  | PROTECT | 6374 (83.99%) | 1022 (13.47%) | 116 (1.53%) | 77 (1.01%) |
|  |  |  | **Total** | **252910 (79.32%)** | **50049 (15.70%)** | **8978 (2.82%)** | **6926 (2.17%)** |
| Depressed Mood | Feeling down, depressed, or hopeless | 20510/120105 | UK Biobank (MHQ) | 112187 (78.36%) | 26516 (18.52%) | 2690 (1.88%) | 1778 (1.24%) |
|  |  |  | UK Biobank (EoP) | 119004 (77.81%) | 28914 (18.91%) | 2998 (1.96%) | 2016 (1.32%) |
|  |  |  | GLAD | 3595 (23.70%) | 6746 (44.47%) | 2538 (16.73%) | 2292 (15.11%) |
|  |  |  | PROTECT | 6108 (80.48%) | 1329 (17.51%) | 92 (1.21%) | 60 (0.79%) |
|  |  |  | **Total** | **240894 (75.55%)** | **63505 (19.92%)** | **8318 (2.61%)** | **6146 (1.93%)** |
| Fatigue | Feeling tired or having little energy | 20519/120107 | UK Biobank (MHQ) | 71903 (50.39%) | 56170 (39.23%) | 7595 (5.30%) | 7503 (5.24%) |
|  |  |  | UK Biobank (EoP) | 74733 (48.87%) | 61571 (40.26%) | 8820 (5.77%) | 7808 (5.11%) |
|  |  |  | GLAD | 1335 (8.80%) | 4971 (32.77%) | 3530 (23.27%) | 5335 (35.17%) |
|  |  |  | PROTECT | 3715 (48.95%) | 3164 (41.69%) | 359 (4.73%) | 351 (4.63%) |
|  |  |  | **Total** | **151686 (47.57%)** | **125876 (39.48%)** | **20304 (6.37%)** | **20997 (6.58%)** |
| Feelings of Inadequacy | Feeling bad about yourself – or that you are a failure or have let yourself or your family down | 20507/120109 | UK Biobank (MHQ) | 116091 (81.09%) | 21503 (15.02%) | 2878 (2.01%) | 2699 (1.89%) |
|  |  |  | UK Biobank (EoP) | 123702 (80.89%) | 22756 (14.88%) | 3399 (2.22%) | 3075 (2.01%) |
|  |  |  | GLAD | 3986 (26.27%) | 5131 (33.82%) | 2800 (18.46%) | 3254 (21.45%) |
|  |  |  | PROTECT | 6095 (80.31%) | 1260 (16.60%) | 128 (1.69%) | 106 (1.40%) |
|  |  |  | **Total** | **249874 (78.36%)** | **50650 (15.88%)** | **9205 (2.89%)** | **9134 (2.86%)** |
| Psychomotor Changes | Moving or speaking so slowly that other people could have noticed, or the opposite – being so fidgety or restless that you have been moving around a lot more than usual | 20518/120111 | UK Biobank (MHQ) | 135592 (94.71%) | 5805 (4.05%) | 1025 (0.72%) | 749 (0.52%) |
|  |  |  | UK Biobank (EoP) | 141662 (92.63%) | 8285 (5.42%) | 1676 (1.10%) | 1309 (0.86%) |
|  |  |  | GLAD | 9932 (65.47%) | 3254 (21.45%) | 1215 (8.01%) | 770 (5.08%) |
|  |  |  | PROTECT | 7363 (97.02%) | 182 (2.40%) | 22 (0.29%) | 22 (0.29%) |
|  |  |  | **Total** | **294549 (92.37%)** | **17526 (5.5%)** | **3938 (1.24%)** | **2850 (0.89%)** |
| Sleep Problems | Trouble falling asleep or staying asleep, or sleeping too much | 20517/120106 | UK Biobank (MHQ) | 73219 (51.14%) | 48936 (34.18%) | 10007 (6.99%) | 11009 (7.69%) |
|  |  |  | UK Biobank (EoP) | 70799 (46.29%) | 56955 (37.24%) | 12245 (8.01%) | 12933 (8.46%) |
|  |  |  | GLAD | 2577 (16.99%) | 5043 (33.24%) | 3173 (20.91%) | 4378 (28.86%) |
|  |  |  | PROTECT | 3708 (48.86%) | 2749 (36.22%) | 570 (7.51%) | 562 (7.41%) |
|  |  |  | **Total** | **150303 (47.14%)** | **113683 (35.65%)** | **25995 (8.15%)** | **28882 (9.06%)** |
| Suicidal Thoughts | Thoughts you would be better off dead or of hurting yourself in some way | 20513/120112 | UK Biobank (MHQ) | 137381 (95.96%) | 4801 (3.35%) | 566 (0.40%) | 423 (0.30%) |
|  |  |  | UK Biobank (EoP) | 146843 (96.02%) | 4880 (3.19%) | 686 (0.45%) | 523 (0.34%) |
|  |  |  | GLAD | 9116 (60.09%) | 3713 (24.47%) | 1215 (8.01%) | 1127 (7.43%) |
|  |  |  | PROTECT | 7392 (97.40%) | 172 (2.27%) | 13 (0.17%) | 12 (0.16%) |
|  |  |  | **Total** | **300732 (94.31%)** | **13566 (4.25%)** | **2480 (0.78%)** | **2085 (0.65%)** |

#### 2.1.2 Study population

In each GWAS sample, individuals were only retained if they had reported European ancestry and had provided a valid response to all PHQ-9 items. Individuals were excluded if they had reported a previous professional diagnosis of schizophrenia, psychosis, mania, hypomania, bipolar, or manic depression (UKB Field ID: 20544) or a previous prescription of medication for a psychotic experience (UKB Field ID: 20466).

#### 2.1.3 GWAS software

All GWAS were conducted using REGENIE v3.1.3^48^. In step one of REGENIE, ridge regression is applied to a subset of quality controlled (QC’d) variants to fit, combine and decompose a set of leave-one-chromosome-out (LOCO) predictions. QC for step one was undertaken using PLINK v1.9^49^. In step two, imputed variants are tested for association with the phenotype. LOCO predictions from step one are included as covariates to control for proximal contamination. For all GWAS, genotyping batch, sex, age, and age-squared were included as covariates, as were the maximum available genetic principal components (PCs) for GLAD (10 PCs) and PROTECT (20 PCs) to control for population stratification. For the UK Biobank analyses, 16 PCs were included as recommended by Privé et al.^50^. Assessment centre was also included as a covariate for UK Biobank analyses.

A total of 40 GWAS were conducted for the meta-analyses – one for each of the nine PHQ-9 depression symptom phenotypes as well as the sum-score across all nine items in each of the four samples. To maximise statistical power, PHQ-9 phenotypes were treated as continuous (ranging 0-3 for individual items and 0-27 for the sum score) and analysed using linear regression. GWAS were restricted to the autosomes.

#### 2.1.4 GWAS: UK Biobank (UKB)

UKB is a large-scale biomedical database and research resource consisting of ∼500,000 individuals with data across a broad range of phenotypes, including mental health outcomes^44^. Individuals in UKB have been genotyped on the custom UK Biobank Axiom or UKBiLEVE arrays, with imputed data available for ∼90 million variants imputed with IMPUTE2 using the Haplotype Reference Consortium (HRC)^51^ and combined UK10K + 1000 Genomes Phase 3 reference panels^52^.

UKB participants have completed the PHQ-9 in two online surveys. In total, 157,345 individuals provided responses as part of the Mental Health Questionnaire (UKB-MHQ) (Category: 136) between 2016 and 2017, and 167,199 individuals had provided responses as part of the Experience of Pain Questionnaire (UKB-EoP) (Category: 154) between 2019 and 2020.

After filtering for self-reported European ancestry, valid PHQ-9 responses, and previous diagnosis/prescription exclusions, 144,630 (UKB-MHQ) and 155,027 (UKB-EoP) individuals remained prior to genetic QC for REGENIE. In step one, single-nucleotide polymorphisms (SNPs) with a call rate > 98%, minor allele frequency (MAF) > 1%, and Hardy-Weinberg equilibrium test *p* > 1x10^-8^ were retained, as were individuals with variant missingness < 2%, no unusual levels of heterozygosity, and not mismatched on sex. Individuals were retained if they were determined to be of European ancestry based on 4-means clustering on the first two principal components.

For the final GWAS analyses, 143,171 (mean age [SD] = 63.70 [7.68]; %female = 56.38%) and 152,932 (mean age [SD] = 65.95 [7.63]; %female = 56.57%) individuals proceeded from the MHQ and EoP questionnaires respectively. Of these, 108,601 individuals had provided responses on both questionnaires. In step two, a total of 9,746,698 imputed variants were retained with MAF ≥ 0.01 and imputation INFO score ≥ 0.7.

#### 2.1.5 GWAS: Genetic Links to Anxiety and Depression (GLAD) Study

The GLAD study has the specific goal of recruiting a large cohort of re-contactable individuals with anxiety or depression into the National Institute for Health and Care Research (NIHR) Mental Health BioResource with genetic, environmental and phenotypic data collected^41^. Genotyping for GLAD was conducted using the UK Biobank v2 Axiom array and imputed using the TopMed imputation pipeline^53^.

After filtering for self-reported European ancestry, valid PHQ-9 responses, and previous diagnosis/prescription exclusions, 15,472 individuals remained prior to genetic QC for REGENIE step one. Genotype data was provided by the study team and had been filtered to retain SNPs with a genotype call rate > 95%, MAF > 1%, Hardy-Weinberg equilibrium test *p* > 1x10^-10^, and individuals with genotype missingness < 5%. Individuals were additionally excluded if they had unusual levels of heterozygosity, mismatched on sex and of non-European ancestry based on 4-means clustering. A total of 15,171 individuals (mean age [SD] = 39.27 [14.61]; %female = 78.30%) were retained for the final analysis. In step two, a total of 13,979,187 imputed variants with MAF ≥ 0.001 and INFO ≥ 0.7 were analysed.

#### 2.1.6 GWAS: PROTECT Study

PROTECT is an online registry of ∼25,000 UK-based individuals that aims to track cognitive health in older adults. Individuals were only considered eligible for inclusion in PROTECT if they were older than 50, had no previous dementia diagnosis and had internet access. Genetic data are available alongside phenotypic data for ∼10,000 of the participants. These individuals were genotyped on the Illumina Infinium Global Screening Array and imputed on the 1000 Genomes reference panel^54^ using the Michigan imputation server and genotype phasing using Eagle.

After filtering for self-reported European ancestry, valid PHQ-9 responses, and previous diagnosis/prescription exclusions, 7,589 individuals remained for genetic QC for step one of REGENIE. Genetic data in PROTECT had been previously QC’d prior to imputation to only retain individuals and variants with a call rate > 98%, Hardy-Weinberg equilibrium test *p* > 0.00001 and excluding unusual heterozygosity^42^. Variants used in step one were down-sampled from the imputed data using a snplist from the Illumina Infinium Global Screening Array provided by the PROTECT investigators. Variants were retained if they had MAF > 1%. After mismatched sex and 4-means clustering ancestry exclusions, a total of 7,589 individuals (mean age [SD] = 61.96 [7.07]; %female = 75.13%) proceeded to step two. In step two, 9,388,534 imputed variants with MAF ≥ 0.001 and imputation INFO score ≥ 0.7 were analysed.

### 2.2 GWAS summary statistics

An overview of additional summary statistics obtained for this study can be seen in Table 2.

**Table 2:**
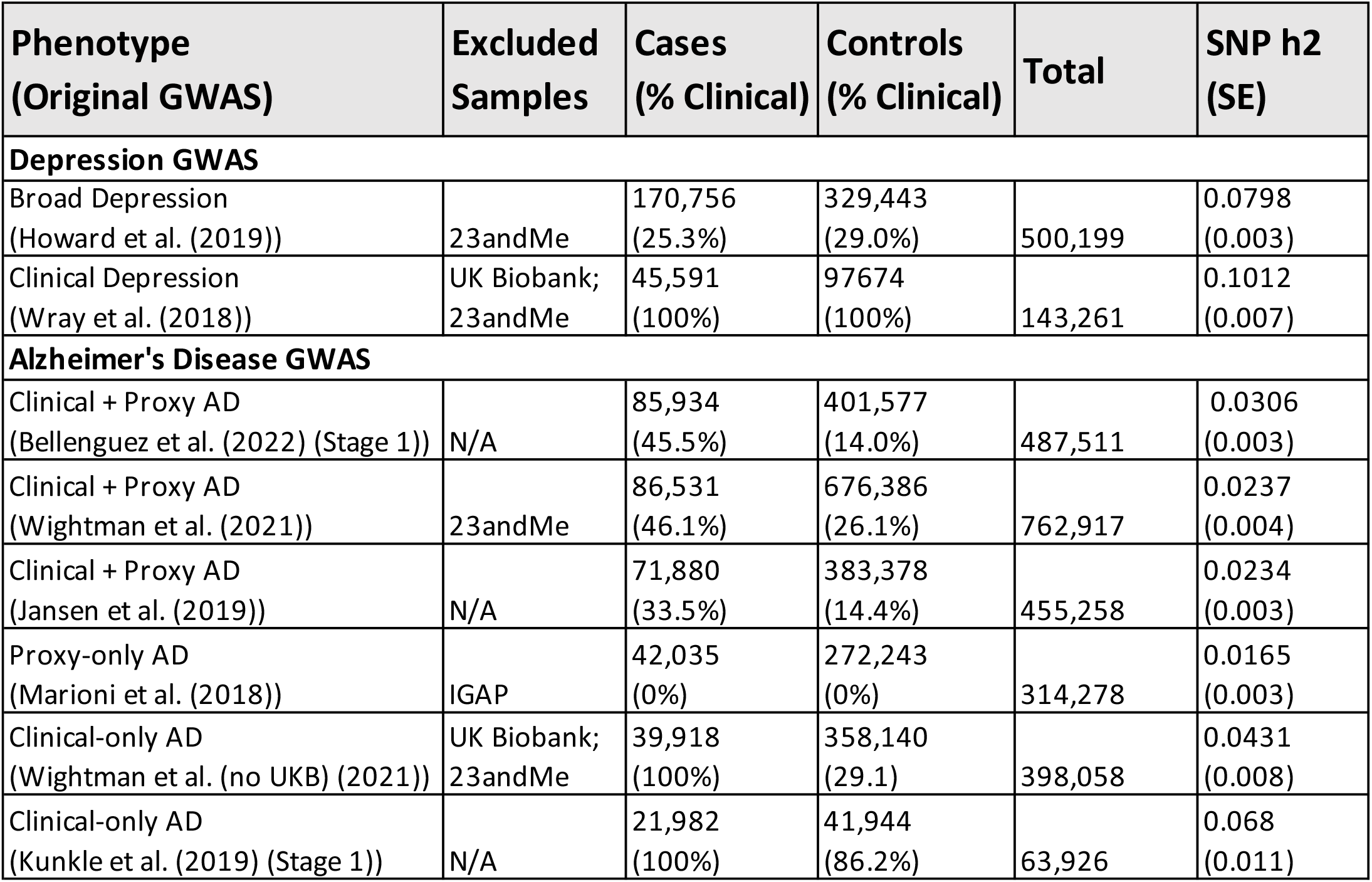
An overview of previously conducted genome-wide association studies for depression and Alzheimer’s disease used in this study. Heritability estimates were calculated naively on the liability scale from these standardised summary statistics using Linkage Disequilibrium Score Regression (LDSC), taking a population prevalence of 15% for depression and 5% for Alzheimer’s disease.

#### 2.2.1 Clinical and broad depression

To examine potential differences in genetic overlap with AD between depression as a disorder compared to individual depression symptoms, summary statistics for two previously conducted GWAS of clinical and broad depression were obtained from the Psychiatric Genomics Consortium (PGC) (https://pgc.unc.edu/for-researchers/download-results/). For clinical depression, we used a subsample of the Major Depressive Disorder (MDD) GWAS by Wray et al.^11^ that excluded samples from the UKB and 23andMe, and contained only individuals for whom case ascertainment was defined through structured diagnostic interview or electronic health records. For the broad definition depression GWAS, we used a subsample of the depression GWAS by Howard et al.^12^, which also excluded samples from 23andMe. In addition to clinical cases and controls used by Wray et al.^11^, this broad depression GWAS included individuals in the UKB for whom case-control ascertainment was based on self-reported responses to the questions “Have you ever seen a general practitioner for nerves, anxiety tension or depression?” and “Have you ever seen a psychiatrist for nerves anxiety, tension or depression?”.

#### 2.2.2 Alzheimer’s disease

Summary statistics were obtained from six previously conducted AD GWAS: three with proxy+clinical, one with proxy-only and two with clinical-only case ascertainment. All three of the proxy+clinical AD GWAS – Bellenguez et al.^17^, Wightman et al.^16^ and Jansen et al.^15^ – and the proxy-only AD GWAS – Marioni et al.^37^ – used data from the UKB for proxy AD samples.

There are some key differences in the way these AD GWAS define proxy cases and controls. Bellenguez et al.^17^ define proxy cases/controls as a binary phenotype, whereby individuals reporting a parent with AD or dementia are considered cases and those reporting no parental history are considered controls. Wightman et al.^16^ and Jansen et al.^15^ instead define proxy cases/control as a continuous phenotype, summing the number of parents an individual has reported with dementia and down-weighting unaffected parents by their age (or age of death).

For the proxy-only Marioni et al.^37^ GWAS, summary statistics were obtained from a meta-analysis of paternal and maternal AD. Here, proxy phenotyping was based on the self-report of either maternal or paternal AD, including the parent’s age at time of reporting/age of death as a covariate.

Summary statistics from a clinical-only subsample of the GWAS by Wightman et al. that excluded proxy cases/controls from the UKB^45^ were obtained from the authors. Summary statistics for a final clinical-only AD GWAS were obtained from Stage 1 of the GWAS by Kunkle et al.^14^.

### 2.3 Summary statistic standardisation

Summary statistics from all 40 depression symptom GWAS, the two depression GWAS and the six AD GWAS, were standardised using the MungeSumstats^55^ in R version 4.2.1. Using dbSNP 141 and the BSgenome.Hsapiens.1000genomes.hs37d5 reference genome, missing rsIDs were corrected, duplicates and multi-allelic variants removed, effect alleles and the direction of their effects aligned to the reference genome, and variants filtered at INFO score ≥ 0.7 and MAF ≥ 0.01. The GLAD Study and Bellenguez et al.^17^ summary statistics were lifted over from GRCh38 to GRCh37.

### 2.4 SNP heritability

SNP heritability (*h^2^_SNP_*) estimates were calculated for all GWAS used in this study with Linkage Disequilibrium Score Regression (LDSC)^56, 57^. Briefly, LDSC calculates *h^2^_SNP_* by regressing the effect sizes from GWAS summary statistics on their LD score as computed in a reference panel – in this case HapMap3 variants contained within the European sample of 1000 Genomes Phase 3. Liability scale *h^2^_SNP_* was calculated naively from the standardised depression GWAS and AD GWAS using a 15% and 5% population prevalence respectively^11, 16^. Heritability *Z*-scores were calculated for all phenotypes by dividing the *h^2^_SNP_* estimates by their standard error.

### 2.5 GWAS meta-analysis of depression symptoms

To leverage the maximum genetic information available controlling for the sample overlap between the UKB-MHQ and UKB-EoP samples, REGENIE output for each PHQ-9 phenotype from the UKB-EoP, GLAD Study and PROTECT were first subject to Inverse Variance Weighted (IVW) meta-analysis using METAL^58^. All available variants were included, for a total of 8,425,618 (*N* = 175,692). Multi-trait Analysis of GWAS (MTAG)^59^ v1.0.8 was then used to meta-analyse the METAL output with the UKB-MHQ sample. While MTAG is commonly used for the joint genetic analysis of multiple traits or multiple measurements of the same trait, by assuming the heritability of included phenotypes are equal (*--equal-h2*) and their genetic correlation is one (*--perfect-gencov*), MTAG performs an IVW meta-analysis of the same measures of the same trait, accounting for sample overlap using the cross-trait intercept from LDSC^56, 57^. Heritability estimates for all samples, plus the METAL meta-analysis, are in Supplementary Table 2. Genetic correlations between the UKB-MHQ and METAL GWAS are in Supplementary Table 3. For greater detail on the IVW function of MTAG, see the online methods of the original MTAG paper^59^. A total of 8,196,874 SNPs with MAF > 0.01 were present for MTAG analysis.

This MTAG function provides one set of summary statistics and two GWAS-equivalent sample sizes – one for each original sample included. A single, weighted GWAS-equivalent *N* was obtained for each PHQ-9-MTAG GWAS, using the following formula;

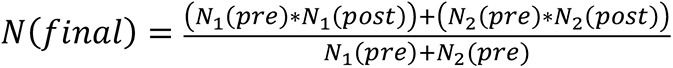

where *N_1_* and *N_2_* represents the UKB-MHQ and METAL GWAS respectively, *pre* is the mean sample size prior to inclusion in MTAG and *post* is the GWAS-equivalent sample estimated by MTAG following analysis.

### 2.6 Genomic risk loci and gene annotation

GWAS meta-analysis results were annotated using FUMA GWAS^60^ v3.1.6a. Genome-wide significance was set at *p* ≤ 5e-8. Lead variants at genomic risk loci were defined by clumping all variants correlated at r^2^ > 0.1 250kb either side. Clumping was performed using the European sample of the 1000 Genomes Phase 3 reference panel. Lead variants were mapped to genes within 10kb using positional mapping and eQTLs from four brain (BrainSeq^61^, PsychENCODE^62^, CommonMind^63^, and BRAINEAC^64^) and five blood (BloodeQTL^65^, BIOS^66^, eQTLGen cis and trans^67^, Twins UK^68^ and xQTLServer^69^) eQTL datasets, alongside all 54 tissue type eQTLs from GTEx v8 (https://gtexportal.org/home/tissueSummaryPage).

### 2.7 Genetic correlations

Genetic correlation can be understood as the genome-wide correlation of genetic effects between two phenotypes, and as such can be viewed as an estimate of pleiotropy^70^.

Genetic correlations were calculated between each depression phenotype and the six AD GWAS using High Definition Likelihood v1.4.1 (HDL)^71^. HDL extends the LDSC framework by leveraging LD information from across the whole LD reference panel through eigen decomposition, thus shrinking standard errors and improving precision. A pre-computed eigenvector/value LD reference panel calculated from 335,265 individuals of European ancestry in the UKB was obtained via the HDL GitHub (https://github.com/zhenin/HDL/wiki/Reference-panels). This reference panel was calculated using 1,029,876, imputed, autosomal HapMap3 SNPs, with bi-allelic SNPs outside the MHC region, MAF > 5%, a call rate > 95% and INFO > 0.9 retained.

### 2.8 Local genetic correlations

Local genetic correlation assesses the correlation of genetic effects between two phenotypes in specific region of the genome. It provides a more refined examination of the genetic overlap between two phenotypes, allowing for the identification of key regions driving shared genetic architecture.

LAVA^72^ was used to assess regions of local genetic correlation between each depression phenotype and the six AD GWAS across 2495 semi-independent, pre- defined LD-blocks of at least 2500 base pairs (https://github.com/josefin-werme/LAVA). Loci-specific heritability estimates were calculated for each phenotype for each block. If both a depression phenotype and an AD GWAS showed significant local heritability at a specific locus (Bonferroni corrected *p*-value ≤ 2e-5 (0.05/2495)), bivariate local genetic correlation was tested. Bivariate results were considered significant at *p*_FDR_ ≤ 0.05 correcting for the total number of bivariate tests. Sample overlap was accounted for using an LDSC intercept matrix. Analysis was restricted to the 5,531,969 non-strand-ambiguous variants shared across all GWAS summary statistics and on the European ancestry 1000 Genomes Phase 3 reference panel.

### 2.9 Colocalisation

Colocalisation using COLOC is a Bayesian statistical method to assess the probability of two phenotypes sharing a causal variant in a pre-defined genomic region.

Within the colocalisation framework, posterior probability is assessed for five hypotheses:

H_0_: There is no causal variant in the region for either phenotype
H_1_: There is a causal variant in the region for the first phenotype
H_2_: There is a causal variant in the region for the second phenotype
H_3_: Distinct causal variants for each trait in the region
H_4_: Both traits share a causal variant.

Colocalisation analysis was conducted using the COLOC-reporter pipeline^73^ (https://github.com/ThomasPSpargo/COLOC-reporter). COLOC-reporter extracts variants in user-defined genomic regions and calculates the LD matrix for this region from a user-defined reference panel. It then harmonises the summary statistics to match the allele order of the reference panel, flipping effect directions accordingly. Observed versus expected Z-scores are assessed using the diagnostic tools provided in the susieR R package^74^. Z-score outliers are omitted. Following this QC, Sum of Single Effects (SuSiE) fine-mapping^74^ is conducted to identify 95% credible sets in these regions for each phenotype. Identification of credible sets in both phenotype allows for the relaxation of the single causal variant assumption. All possible credible sets are then assessed pairwise for shared signal between phenotypes, improving resolution for colocalisation inference in regions containing multiple signals^75^. Should no 95% credible set be identified or only identified for one phenotype, colocalisation under the single causal variant assumption is performed using the *coloc.abf*^76^. A posterior probability ≥ 80% for H_4_ was considered evidence of colocalisation between two phenotypes (PP.H_4_ ≥ 0.8). The SuSiE model assumed at most 10 causal variants (L = 10) per credible set. We used default priors.

Trait pairs with LAVA correlations significant at *p*_FDR_ ≤ 0.05 were passed to COLOC-reporter. Where the same depression phenotype showed nominally significant local genetic correlation at the same locus but with different AD GWAS, these phenotype pairs were included as a sensitivity analysis. Regions +/-250kb (r^2^ ≥ 0.1) from lead variants at genome-wide significant loci from the MTAG-PHQ-9, broad and clinical depression GWAS were also examined for evidence of colocalisation with the 6 AD GWAS.

### 2.10 Mendelian randomisation (MR)

MR is a statistical method that uses genetic variants associated with an exposure as instrumental variables to assess the causal effect of that exposure on an outcome of interest^77^. Two Sample MR framework estimates the causal relationships between an exposure and outcome using GWAS summary statistics. Valid MR instruments are defined by three key assumptions: (1) relevance – IVs are strongly associated with the exposure of interest; (2) independence – there are no confounders in the association between IVs and the outcome of interest; and (3) exclusion restriction – instruments are not associated to the outcome other than via the exposure, for example through horizontal pleiotropy^77^.

Sample overlap is a known source of bias in Two Sample MR^78^. Given the likelihood of sample overlap between the depression phenotypes and the AD GWAS containing proxy cases/controls due to participants from the UKB, MR analysis was primarily conducted using Causal Analysis Using Summary Effect estimates (CAUSE)^79^ v1.2.0. CAUSE is a Bayesian MR method robust to sample overlap, correlated and uncorrelated pleiotropy, also avoiding exclusion restriction assumption violations^80^.

In CAUSE, the presence of a significant causal effect, γ, is derived by comparing the expected log pointwise posterior density (ELPD) of a sharing model with γ fixed to zero to a causal model where γ is a free parameter for a one-tailed p-value test. The posterior median of γ describes the causal effect estimate and is reported alongside its 95% credible intervals. CAUSE uses a larger set of instruments that traditional MR methods. As such, clumping was performed with the default setting at r^2^ ≥ 0.01 and a *p*-value ≤ 0.001 within a 10,000kb window.

Causal estimates were also calculated using the traditional IVW method. IVW estimates are biased by the presence of horizontal pleiotropy^81^. Sensitivity analyses were therefore conducted using MR-Egger, weighted-median and weighted-mode MR. These methods allow for varying degrees of pleiotropy while providing unbiased causal estimates^82–84^. MR-PRESSO^85^ was also implemented. MR-PRESSO identifies and excludes outlying instruments based on their contribution to heterogeneity and provides a corrected causal estimate.

Pleiotropy was assessed using the MR-Egger intercept test^86^ (significant pleiotropy: *p* ≤ 0.05). Heterogeneity tests were also conducted for IVW and MR-Egger estimates using their respective Cochran’s Q test^87^ (significant heterogeneity: *p*-value ≤ 0.05). Instrument strength was calculated via the minimum and mean F-statistic (recommended F-statistic ≥ 10)^88^ (β^2^/SE^2^). I^2^ was calculated to ensure measurement error was sufficiently low so as to ensure validity of results from MR-Egger (recommended I^2^ ≥ 0.90)^89^.

For IVW-MR, MR-Egger, weighted-median and weighted-mode MR, instruments clumped at a r^2^ ≥ 0.001 and a *p*-value ≤ 5e-8 within 10,000kb. Where no instruments were available at *p* ≤ 5e-8 or where < 5 instruments were available, a *p* ≤ 5e-6 threshold was used.

For all analyses, instruments were clumped using data from individuals of European ancestry in 1000 Genomes Phase 3. IVW, MR-Egger, weighted-median and weighted-mode analyses were conducted using the TwoSampleMR package v0.5.6.

The *APOE* gene is known to be associated with non-AD phenotypes such as cardiovascular disease^90^ and type 2 diabetes (T2D)^91^. Both have been linked to depression in previous MR analyses^92, 93^. As such, the inclusion of *APOE* would violate MR’s independence assumption. All MR analyses were therefore conducted excluding variants in the APOE region (chr19:45,020,859–45,844,508 (GRCh37)) as per Lord et al.^94^.

### 2.11 Polygenic risk scores (PRS)

Polygenic risk scores (PRS) describe the sum of an individual’s risk alleles, weighted by their effect size^95^. The ten PHQ-9-MTAG, clinical depression and broad definition depression GWAS were processed for PRS using the BayesR-SS function contained within MegaPRS and implemented in LDAK v5.2.1.^96^. BayesR-SS assumes the BLD-LDAK heritability model, which incorporates 65 genome annotations pertaining to genomic features such as whether the variants are in coding regions or highly conserved^96^. Annotation files were obtained from the LDAK website (http://dougspeed.com/bldldak/). PRS calculation was restricted to the 1,217,311 HapMap3 SNPs, with strand ambiguous SNPs excluded.

PRS predictive utility for AD case/control status was assessed using logistic regression in three clinically ascertained AD cohorts: AddNeuroMed and Dementia Case Register Studies (ANM)^97^ (*N_cases_* = 564; *N_controls_* = 345), the Alzheimer’s Disease Neuroimaging Initiative (ADNI) (https://adni.loni.usc.edu) (*N_cases_* = 356; *N_controls_* = 360), and the Genetic and Environmental Risk in Alzheimer’s Disease (GERAD1) Consortium (https://portal.dementiasplatform.uk/CohortDirectory/Item?fingerPrintID=GERAD) (*N_cases_* = 2,661; *N_controls_* = 1,124) (Supplementary Table 4). All analyses controlled for age, sex, and 10 PCs, and were restricted to individuals aged ≥ 65 to provide clean controls. As a sensitivity analysis, PRS were also calculated in these cohorts excluding the *APOE* region. Genetic QC and imputation steps for these cohorts can be viewed in detail in the study by Lord et al.^98^.

## 3. RESULTS

### 3.1 PHQ-9 genome-wide meta-analyses

The MTAG meta-analyses identified a total of 40 genomic risk loci between the ten PHQ-9 phenotypes (GWAS equivalent *N* range: 224,535 – 308,421). Only one depression symptom – suicidal thoughts – identified no genome-wide significant variants. Three lead SNPs were shared with more than one PHQ-9 phenotype, leaving a total of 37 unique genomic risk loci (Table 3). The significance of each of the lead variants in each of the samples contributing to the meta-analysis can be seen in Supplementary Table 5. eQTL mapping in FUMA mapped lead variants at genomic risk loci to 76 genes (Supplementary Table 6). *h^2^_SNP_* for the MTAG-PHQ-9 GWAS ranged from 1.12% for suicidal thoughts to 6.78% for the PHQ-9 sum-score. *h^2^_SNP_ Z*-scores were all > 4 (range: 6.59 – 18.50) (Supplementary Table 7), indicating sufficient heritability to obtain reliable genetic correlation estimates in downstream analyses^57^. Genomic inflation factors (λ_GC_) ranged from 1.0638 to 1.2156, with LDSC intercepts ranging from 0.9997 to 1.0007, indicating inflation was due to polygenic signal as opposed to confounding due to population stratification^56, 99^. Manhattan and QQ plots can be viewed in Figure 2.

**Table 3:**
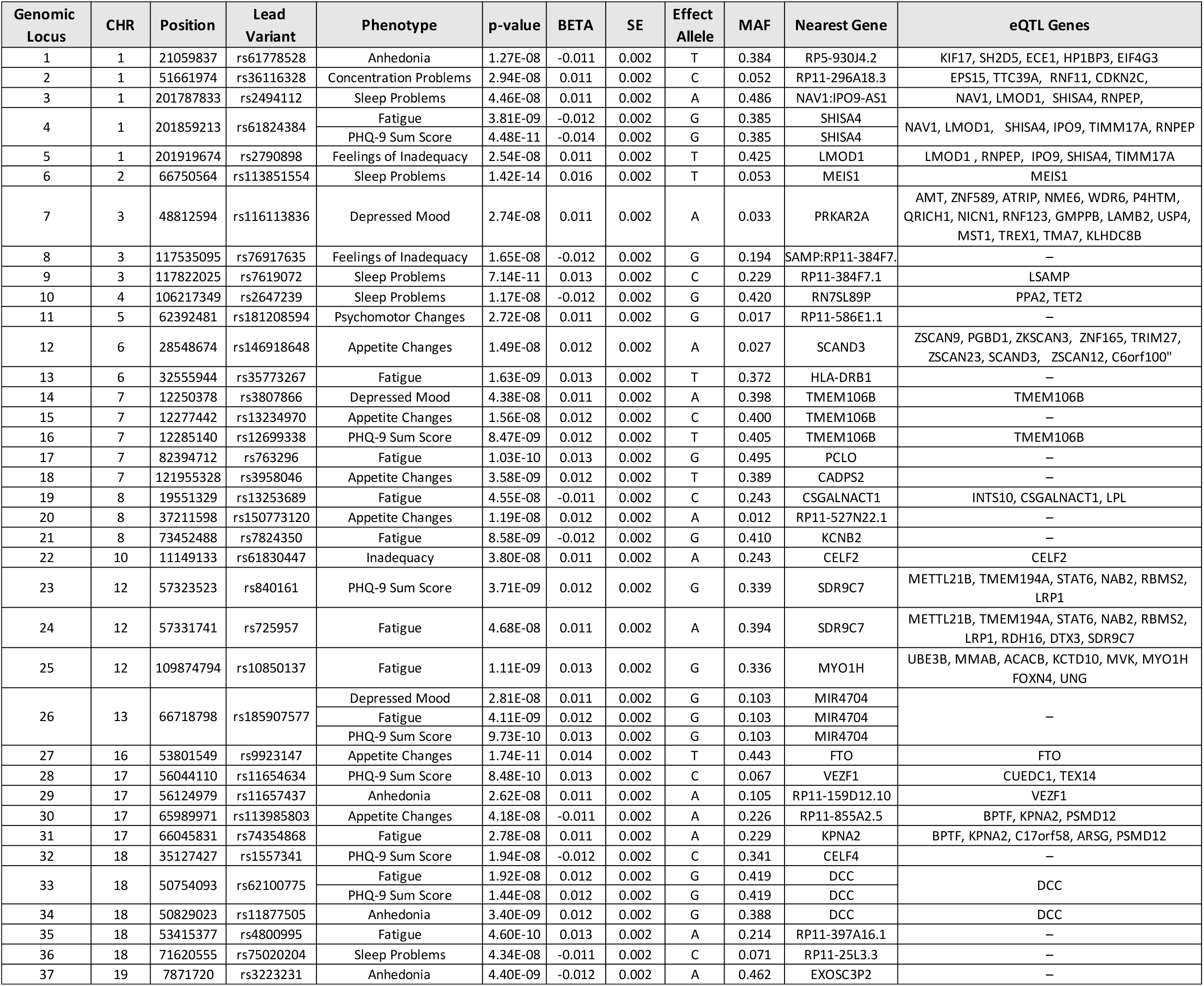
Genomic risk loci from the 10 MTAG-PHQ-9 genome-wide association meta-analyses. Nearest gene is based on positional mapping and ANNOVAR annotation. Expression quantitative trait loci (eQTL) mapping was limited to pFDR ≤ 0.05 in the selected eQTL datasets (see: Methods). Specific tissues with significant pFDR can be seen in Supplementary Table 2.

**Figure 2:**
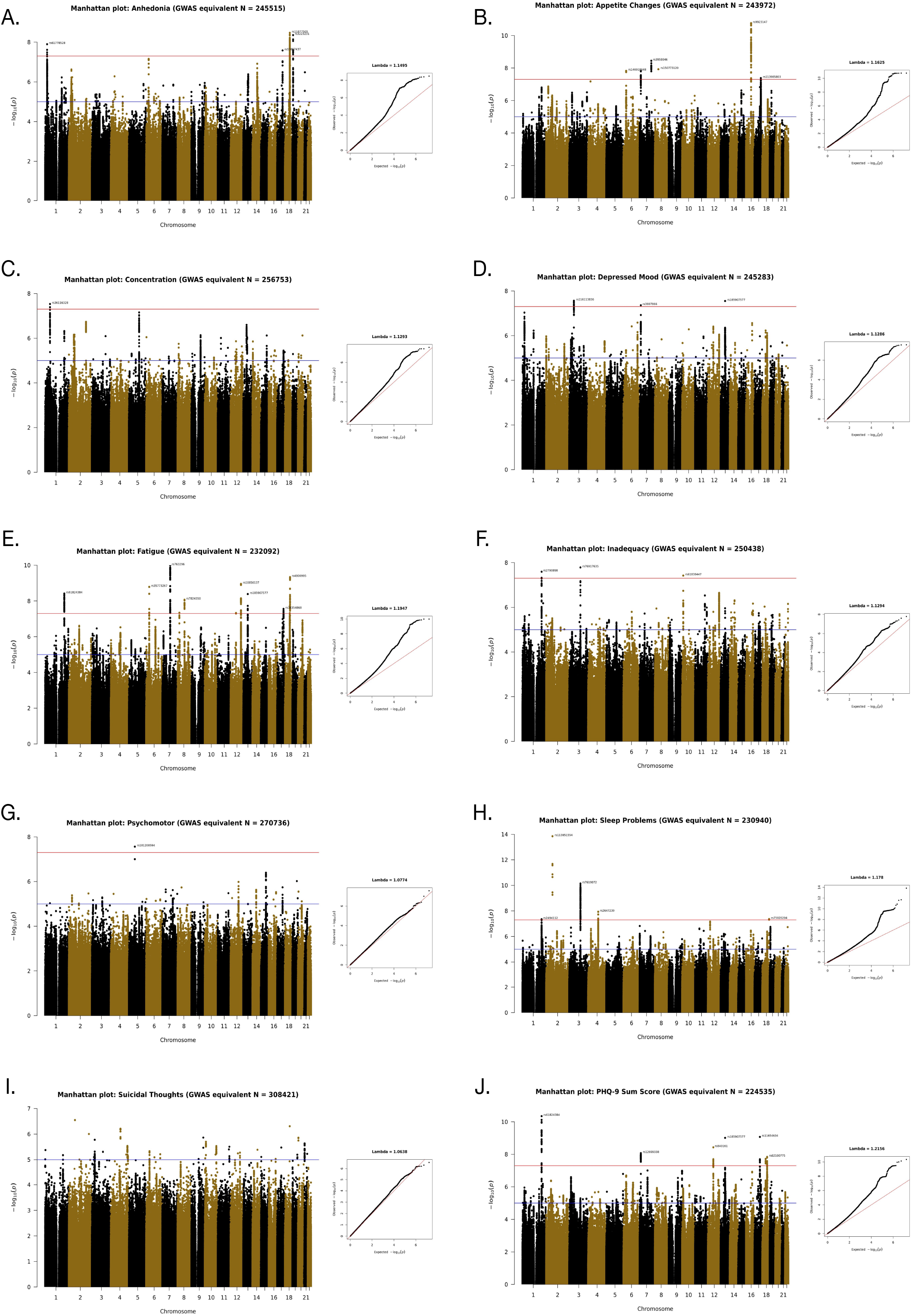
Manhattan and QQ-plots for each of the 10 MTAG-PHQ-9 GWAS meta-analyses. The red lines on the Manhattan plots indicate genome-wide significance (p ≤ 5 x 10^-8^), while the blue lines indicates suggestive significance (p ≤ 1e-5). Top variants at each genomic risk loci are annotated.

### 3.2 Genetic correlations

Of the 72 bivariate genetic correlations calculated between the 12 depression phenotypes and the six AD GWAS, 24 were nominally significant and 20 remained significant at *p*_FDR_ ≤ 0.05 (*r_g_* range: -0.25 – 0.35; *p*-value range: 1.25 x 10^-2^ – 4.01 x 10^-5^; *p*_FDR_ range: 4.5 x 10^-2^ – 1.9 x 10^-3^). Of these, 19 were identified when the AD GWAS in the pair contained either clinical+proxy cases and controls, or proxy-only cases and controls (Figure 3; Supplementary Table 8). Only one *p*_FDR_ significant association was found when using a clinical AD GWAS – between suicidal thoughts and Wightman et al. (*r_g_* = -0.25, p = 6.78 x 10^-3^, *p*-value = 6.78 x 10^-3^; *p*_FDR_ = 3.48 x 10^-2^). All depression phenotypes were significantly genetically correlated with each other (*r_g_* range: 0.57– 0.98; *p*-values ≤ 3.71 x 10^-23^) (Supplementary Table 9, Supplementary Material 1). Only one PHQ-9 symptom pair – concentration problems and psychomotor changes – showed a genetic correlation that was not statistically different from one (95% CI included one), indicating genetic heterogeneity across depression symptoms.

**Figure 3:**
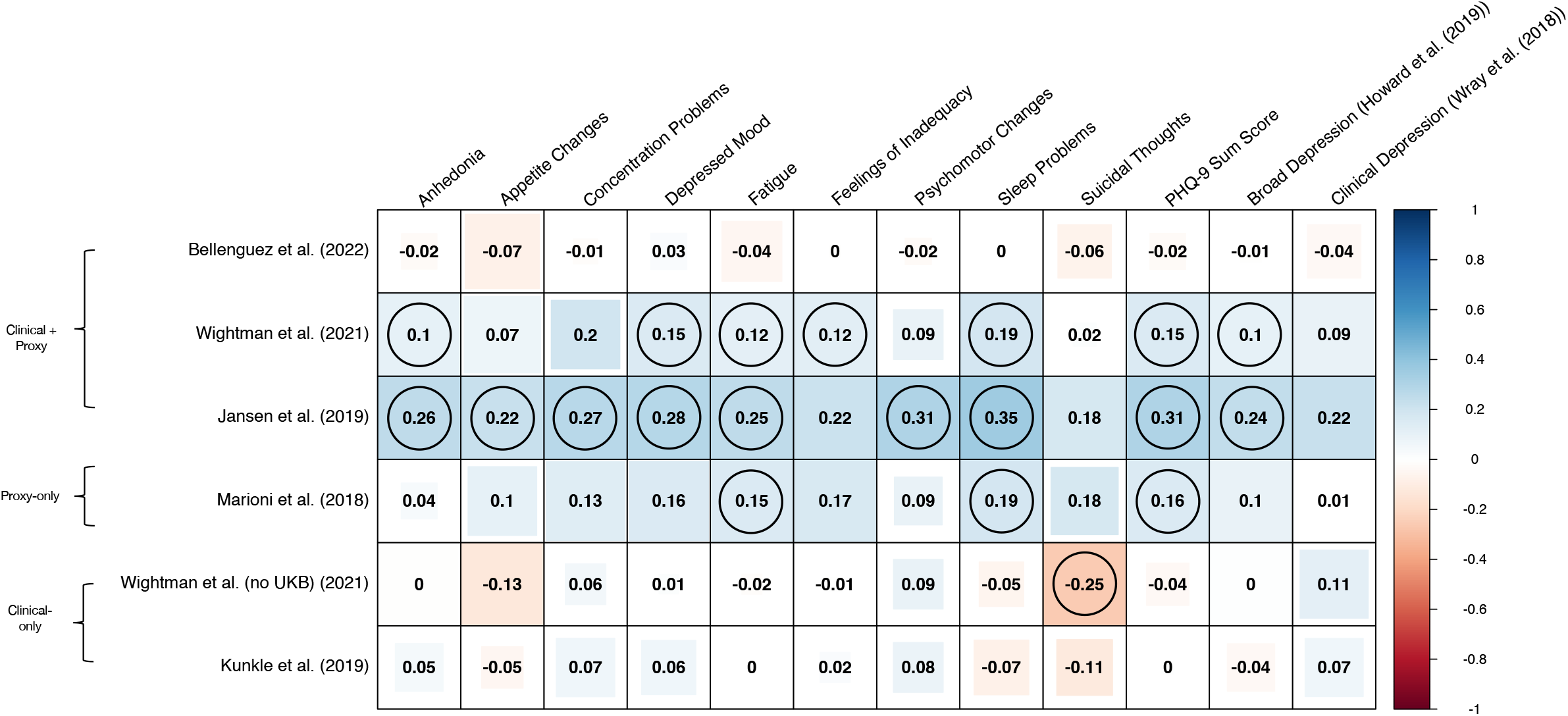
Heatmap of High Definition Likelihood (HDL) calculated genetic correlations between depression items and the six Alzheimer’s disease genome-wide association studies of varying proxy/clinical case/control ascertainment. The colour of each square indicates the strength of the correlation on a scale of -1 to 1. Fill size of the square indicates uncorrected p-value significance. Genetic correlations significant at p_FDR_ ≤ 0.05 are circled.

### 3.3 Local genetic correlations

After univariate testing, a total of 4271 bivariate local genetic correlation tests were conducted across 324 genomic loci. Of these tests, 716 were nominally significant and 15 remained significant at *p*_FDR_ ≤ 0.05 across 14 unique genomic loci (local *r_g_* range: -0.81 – 0.82; *p*-value range: 1.48 x 10^-4^ – 4.2 x 10^-6^; *p*_FDR_ range = 4.22 x 10^-2^ – 1.38 x 10^-2^) (Supplementary Table 10). Of the 15 statistically significant tests, ten were identified when using clinical+proxy/proxy-only AD GWAS. No depression phenotype showed a statistically significant association at the same genomic locus with more than one AD GWAS. However, for ten of the 15 statistically significant tests, nominally significant local genetic correlation was observed between the depression phenotype and at least one additional AD GWAS at the same loci (Supplementary Table 11). Only locus 1790 (chr12:51769420– 53039987) showed *p*_FDR_ significant local genetic correlation with more than one depression phenotype – concentration problems and sleep problems, both with the clinical-only Wightman et al. GWAS. The number of positively and negatively correlated loci identified between each phenotype pair can be viewed in Supplementary Table 12.

### 3.4 Colocalisation

Following LAVA, 14 *p*_FDR_ significant regions of local genetic correlation were passed to the COLOC-reporter pipeline across 15 depression-AD phenotype pairs. A further 14 colocalisation tests were conducted where nominally significant local genetic correlation was observed at a *p*_FDR_ significant loci between the same depression phenotype and a different AD GWAS. As such, a total of 29 statistical colocalisation tests were conducted to follow up LAVA results. No 95% credible sets were identified by SuSiE for any phenotype pairs in these regions. As such, all analyses were conducted under the single causal variant assumption of *coloc.abf*. There was no evidence of colocalisation in any of these loci (mean PP.H_4_ = 0.59%) (Supplementary Table 11). All but two of these tests indicated no causal variant present in either phenotype (PP.H_0_ > 0.8). The two tests in locus 319 (chr2:126754028-127895644), indicated strong probability of a causal variant for the Kunkle et al.^14^ and Wightman et al.^45^ clinical AD GWAS (PP.H_2_ > 0.9) .This locus contains *BIN1*, a known risk gene for AD involved in tau regulation^100, 101^.

An additional 762 colocalisation tests were conducted with the six AD GWAS using regions +/-250kb (r^2^ > 0.1) lead variants from the MTAG-PHQ-9, broad and clinical depression GWAS. SuSiE identified evidence of colocalisation in regions +/- 250kb of lead variants at genomic risk loci 14 (depressed mood), 15 (appetite change), 16 (PHQ-9 sum score) and for broad depression at chr7:12000402-12500402 (PP.H_4_ range: 0.79 – 0.85), all with the same three AD GWAS – Bellenguez et al.^17^, Wightman et al.^16^, and Wightman et al. (excluding the UKB)^45^ (Supplementary Table 13). These colocalisations were all in the region of the transmembrane protein 106B gene (*TMEM106B*) – visualised using LocusZoom^102^ in Figure 4. Colocalisation was also identified for the same phenotype pairs at the same loci under the single causal variant assumption of *coloc.abf* (Supplementary Table 14). The same depression phenotypes and loci were suggestive of colocalisation with Jansen et al.^15^ (PP.H_4_ > 0.6). In a follow-up analysis, we assessed statistical colocalisation +/-250kb *TMEM106B* (chr7:12000920-12532993) between these four AD GWAS and all remaining depression phenotypes. Additional evidence of colocalisation was identified at *TMEM106B* between fatigue and the Bellenguez et al.^17^, Wightman et al.^16^, and Wightman et al. (excluding the UKB)^45^ AD GWAS (Supplementary Table 15), and was suggestive for Jansen et al.^15^ (Supplementary Table 16). Evidence of colocalisation was also suggestive for psychomotor changes with Bellenguez et al.^17^, Wightman et al.^16^, and Wightman et al. (excluding the UKB)^45^ (PP.H_4_ > 0.6).

**Figure 4:**
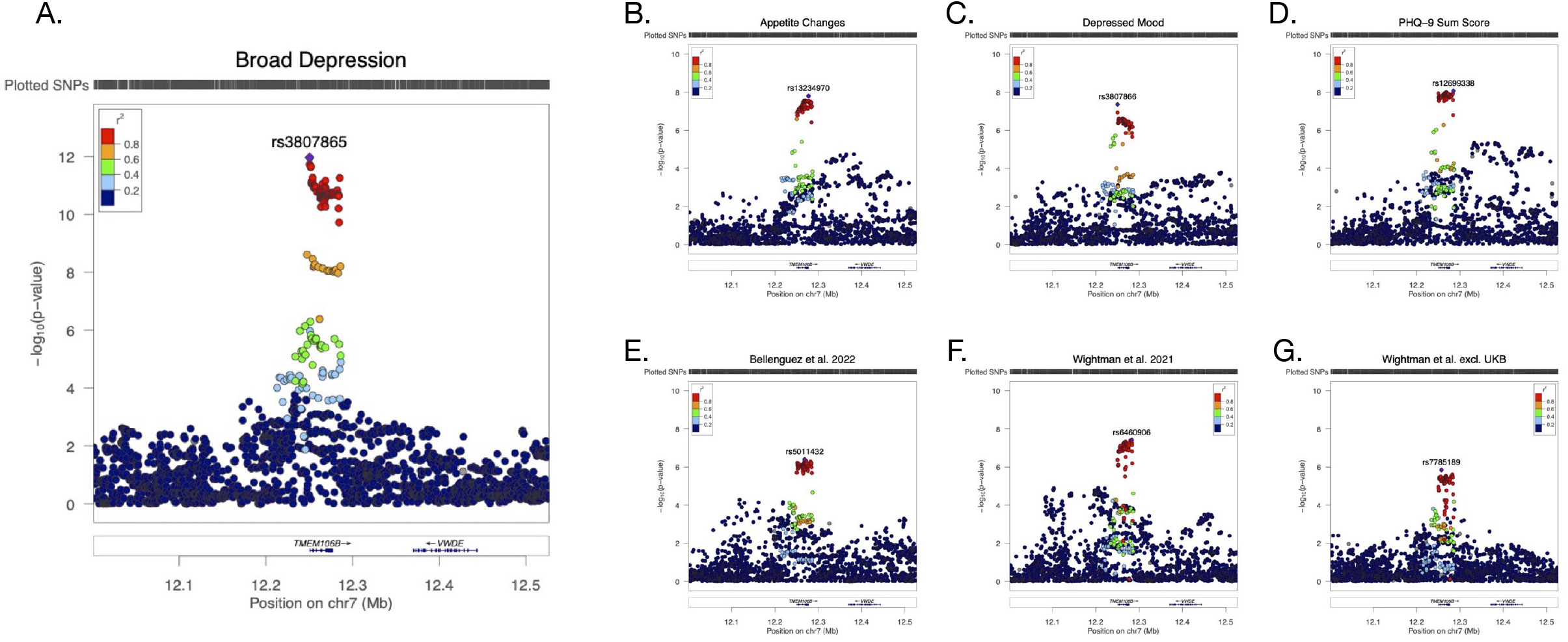
LocusZoom plots of the transmembrane protein 106B (TMEM106B) gene region containing evidence of colocalisation (PP.H_4_ ≤ 0.8). for (A.) broad depression, (B.) appetite changes, (C.) depressed mood, (D.), the PHQ-9 sum score, (E.) Bellenguez et al., (F.) Wightman et al. and (G.) Wightman et al. excluding the UK Biobank. The most significant variant for each phenotype is labelled in their respective plot.

### 3.5 Mendelian Randomisation

We conducted 144 MR tests between the depression phenotypes and AD – 72 in each direction. In CAUSE, no significant causal association was identified between any of the depression items and AD in either direction even at nominal significance (Supplementary Table 17).

F-statistics indicated that instrument strength was sufficient (F_Mean_ range: 22.43 – 63.36; F_Min_ range: 20.84 – 31.56; F_Max_ range: 26.37 – 402.86), as did I^2^ for instrument suitability for MR-Egger (I^2^ range: 0.91 – 0.98). A *p*_FDR_ ≤ 0.05 was applied in each of the other MR methods to correct for the 144 tests conducted, after which no statistically significant associations were observed in any method. Three nominally significant causal associations where a depression item was the exposure (β range = 0.21 – 1.59; *p*-value range = 0.01 – 2e-3) and two where an AD GWAS was the exposure (β range = 0.018 – 0.02; *p*-value range = 0.01 – 0.02) were detected by IVW-MR, of which three we nominally significant in weighted-median MR. No significant pleiotropy (MR-Egger intercept test *p*-value ≤ 0.05) or heterogeneity (IVW Cochran’s Q *p*-value ≤ 0.05) was detected for these five tests. However, nominally significant estimates were only observed between exposures and outcomes where the AD GWAS contained proxy+clinical or proxy-only cases/controls and where the GWAS used for both exposure and outcome were either majority or entirety derived from the UK Biobank, suggesting sample overlap as a potential source of bias. MR-PRESSO detected and excluded significant outliers in 42 tests, returning outlier-corrected causal estimates. Results remained non-significant. Full MR results can be viewed in Supplementary Table 18.

### 3.6 Polygenic risk scores (PRS)

No statistically significant associations were detected between any of the depression phenotype PRS and AD case/control status in any of the three AD target samples (*p*_FDR_ ≤ 0.05, corrected within each sample), with only the suicidal thoughts PRS negatively nominally associated with AD case/control in the ADNI cohort (Nagelkerke’s pseudo-r2 = 0.01, OR [95% CI] = 0.83 [0.72–0.94], β = -0.19, SE = 0.07, *p*-value = 7.90 x 10^-3^). Exclusion of the APOE region had no effect on results (Supplementary Table 19, Supplementary Material 2).

## 4. Discussion

In this study, we performed the first and largest genome-wide meta-analysis of PHQ-9 depression symptom items to date (GWAS equivalent *N* range: 224,535 – 308,421), identifying 37 genomic risk loci. Follow-up analyses examining the genetic overlap between depression/depression symptoms and AD identified 20 significant global correlations and 15 significant local correlations at 14 loci across six AD GWAS with varying proportion of clinical case/control ascertainment. Significant global genetic correlation were primarily with AD GWAS containing proxy cases and controls. No colocalisation was identified at any of the regions of local genetic correlation. However, there was strong evidence of colocalisation between depression several phenotypes and AD in the region of *TMEM106B*. Overall, MR did not suggest that depression or its symptoms were causal for AD, nor vice versa, while polygenic risk scores for depression phenotypes were not predictive of AD case/control status in three clinical AD samples.

The increased power of our PHQ-9 GWAS allowed for the identification of 28 more genomic risk loci than the previous analysis by Thorp et al.^29^. Several of the loci identified in this study have shown previous associations with related phenotypes. For example, *SHISA4* – here identified in association with fatigue symptoms – has been previously implicated as playing a role in disrupted sleep^103^ and daytime napping^104^. The top variant for sleep problems at genomic risk loci 6 (*MEIS1*) – rs113851554 (chr2: 66750564) – was also the top variant in a previous GWAS of insomnia and restless leg syndrome^105^. Additionally, the obesity gene *FTO*^106^ was identified as a genomic risk loci for appetite changes. Although the role of *FTO* in depression is inconclusive^107^, it has been recently linked to anxiety and depression symptoms in individuals with anorexia nervosa (AN)^108^. Consequently, its identification in association with appetite change symptoms – a phenotype relevant to eating behaviours – suggests that symptom-based genetic analysis can help identify the phenotype-relevant biology of individual depression symptoms. Taken alongside the fact that only three of the genomic risk loci were shared between more than one depression symptom phenotype, our GWAS meta-analysis backs findings by Thorp et al.^29^ of genetic heterogeneity between depression symptoms.

Our findings also highlight genetic similarities between depression symptoms. For example, *TMEM106B* – a gene identified in previous depression GWAS^11, 12^ – was the nearest gene to lead variants for three PHQ-9 items – appetite changes (rs13234970), depressed mood (rs3807866) and the PHQ-9 sum score (rs12699338). *TMEM106B* was strongly suggested as a causal gene in a recent multi-ancestry depression GWAS^109^. Further, dysregulation of *TMEM106B* expression has been implicated in association with MDD^110^ and with anxious and weight gain MDD subtypes – both associated with treatment resistance^111^. *TMEM106B* has also been implicated in self-reported diagnosis of anxiety disorder^112^, neuroticism^113^ and in a latent factor GWAS of depressive, manic and psychotic symptoms/disorders^114^, suggesting it is linked to psychiatric risk more generally.

That colocalisation is observed in the *TMEM106B* region between multiple depression phenotypes and both proxy+clinical and clinical-only AD is therefore of particular interest. *TMEM106B* plays a role in lysosomal function – particularly in motor neurons^115^ – and is classically considered a risk gene for frontotemporal dementia^116^. It is has also been identified in recent AD GWAS^16, 17^, and is associated with brain aging, cognitive decline, and neurodegeneration across other brain disorders including Amyotrophic Lateral Sclerosis, Multiple Sclerosis and Parkinson’s Disease^117–122^. Further, *TMEM106B* has been linked to higher CSF levels of neurofilament light (NfL)^123^ – itself predictive of cognitive decline, brain atrophy and cortical amyloid burden in individuals with AD and Mild Cognitive Impairment (MCI)^124^. A recent study also identified higher levels of plasma NfL in individuals with depression, although the sample size of this study was relatively small^125^. Thus, colocalisation between depression phenotypes and AD at *TMEM106B* indicates that depression may itself be genetically linked to overall brain health and resulting general dementia risk. Two previous studies have identified *TMEM106B* as playing a role in both depression and AD^20, 21^. However, our study suggests that this overlap may be driven by the genetic architecture of specific depression symptoms, highlighting the benefits of symptom-level genetic analysis.

A mixed picture is painted across our other downstream analyses. Depression/depression symptom PRS were not predictive of AD case/control status in three clinical samples. Additionally, we do not find evidence of any causal associations using MR. These MR findings are in line with a number of previous studies^19, 39^, but in contradiction to that conducted by Harerimana et al.^22^. In this study, we do not recapitulate the causal association identified by Harerimana et al.^22^ for broad depression on AD as measured by Jansen et al.^15^ – even at a nominal level. This is possibly due to the exclusion of rare variants (MAF < 0.01), and as such future investigation into the role of rare variants in shared risk for depression and AD is warranted. However, the lack of evidence in these analyses versus the observed relationship in epidemiological studies suggests the presence of unidentified confounding. The investigation of possible confounding is an important step for future research to help us better understanded the association between depression and dementia.

While previous studies have shown that the direction of effect in MR can change depending on whether the outcome AD GWAS contains proxy or clinical cases/controls^38, 39^, this study is – to the best of our knowledge – the first to demonstrate a similar effect with genetic correlations. Of the significant genetic correlations we identified, 95% were identified in proxy+clinical or proxy-only AD GWAS. Where two previous studies^20, 22^ identified a genetic correlation between depression and AD, it is noticeable that they used the Jansen et al. proxy+clinial AD GWAS as their primary outcome. Further, in the study by Harerimana et al.^22^ sensitivity analysis did not observe a significant genetic correlation using the clinical-only GWAS by Kunkle et al.^14^.

Exactly why depression/depression symptoms show differences in genetic correlation between proxy and clinical AD is a matter of interest. It is noted that most individuals with dementia are cared for by a family member while they are still living in the community^126^. Caregivers incur considerable lifestyle changes, emotional stress, and social strain^127, 128^ and depression is reported in up to 50% of caregivers for individuals with dementia^129^. As such, the genetic correlations between depression/depression symptoms and proxy-AD may result from increased depression risk due to caregiver responsibilities, exacerbated by a genetic risk. However, this would not explain why no genetic correlations were identified with the Bellenguez et al.^17^ GWAS despite this study also containing proxy+clinical phenotyping. As mentioned, Bellenguez et al.^17^ define proxy cases/controls as a binary phenotype, whereas Wightman et al.^16^ and Jansen et al.^15^ define proxy cases/controls as a continuous phenotype. These phenotyping differences likely partially explain the differences in the genetic correlation results, given that these AD GWAS all use the same data from the UK Biobank. However, the proxy-only Marioni et al.^37^ GWAS – with which genetic correlations were also observed – is a meta-analysis of maternal and paternal AD GWAS where parental age-at-diagnosis/age-of-death is controlled for in the GWAS model, instead of being used for weighting the proxy phenotype prior analysis. Taking this into account, and considering that depression is itself associated with all-cause mortality^130^, it is possible that including age-at-diagnosis/age-of-death in proxy-AD phenotyping induces a form of bias in later cross-trait analyses when the other trait is itself associated with longevity. Further investigation of this issue is required.

Nonetheless, conflicting results such as these pose a problem to researchers seeking to identify genetic relationships between AD and its risk factors. Large differences in the presence or direction of effects depending on which AD GWAS is used to assess associations increases the difficulty in discerning true associations for the purpose of designing interventions. As such, we suggest that future genetic studies examining genetic overlap between AD risk factors and AD conduct primary analyses using a clinically ascertained AD phenotype. A range of AD GWAS with different proxy/clinical ascertainment could then be used to examine the consistency of results in relation to clinical AD. While this approach would limit researchers to AD GWAS with slightly smaller sample sizes for primary analyses, it would also ensure that results are driven not by AD proxy phenotyping alone.

This study has several limitations. Despite being the largest meta-analysis of PHQ-9 items to date, the ability to detect genome-wide significant variants was likely limited by small sample sizes relative to other psychiatric conditions. This includes depression itself, where sample sizes have exceeded one million^13^. Larger samples are required for future studies if we are to truly elucidate the unique genetic architecture of individual depression symptoms. Additionally, our analyses were restricted to individuals of European ancestry. GWAS results may therefore have poor transferability to other ancestry groups. Although efforts are underway to address the issue of lack of diversity in human genetic studies, urgency is required so that studies can be conducted across ancestry groups using equivalent sample sizes. Our study makes also heavy use of data from the UKB. The UKB is known to be affected by healthy volunteer bias, and as a consequence is not fully representative of the wider population^131^.

It is worth noting that this study focused on depression as a risk factor for AD. However, there is evidence that some forms of later life depression are in fact a prodromal phase of dementia onset^8, 132^, and may be related to levels of dementia biomarkers such as plasma Aβ42 and CSF p-tau^133^. As such, dementia-related depression may be biologically distinct from depression as a mental health disorder. While sufficient sample sizes are likely difficult to obtain, future large scale genomic studies of dementia-related depression – as has been undertaken with psychosis in AD^134^ – would prove illuminating on this matter.

In conclusion, this study describes the largest genome-wide meta-analysis of PHQ-9 depression symptom items to date (GWAS equivalent *N* range: 224,535 – 308,421), identifying 37 unique genomic risk loci. Genetic correlations between depression/depression symptoms and AD were primarily observed when the AD GWAS contained clinical+proxy or proxy-only AD case/control ascertainment. Further, this study does not support depression or its symptoms as being causal for AD. However, colocalisation in the *TMEM106B* region between four depression phenotypes and AD across both proxy and clinical AD GWAS suggests future research into the shared biological mechanisms underlying the role of this locus in depression and AD are warranted.

## Data Availability

All GWAS summary statistics generated in the process of conducting this study will be deposited in the NHGRI-EBI Catalog of human genome-wide association studies (GWAS Catalog) following peer-review and publication. This study was pre-registered on the Open Science Framework (https://osf.io/94q35/?view_only=e77f72d4100d47eea7f3ef07dfa9c059).

## 5. Funding and Acknowledgements

LG is funded by the King’s College London DRIVE-Health Centre for Doctoral Training and the Perron Institute for Neurological and Translational. PP is funded by Alzheimer’s Research UK. SK is funded by MSWA and the Perron Institute. HLD acknowledges funding from the Economic and Social Research Council (ESRC). DMH is supported by a Sir Henry Wellcome Postdoctoral Fellowship (Reference 213674/Z/18/Z).

This paper represents independent research part-funded by the NIHR Maudsley Biomedical Research Centre at South London and Maudsley NHS Foundation Trust and King’s College London. The views expressed are those of the author(s) and not necessarily those of the NIHR or the Department of Health and Social Care.

This research has been conducted using the UK Biobank Resource under Application Number 18177. We thank the UK Biobank Team for collecting the data and making it available. We also thank the UK Biobank participants.

We thank the GLAD Study volunteers for their participation, and gratefully acknowledge the NIHR BioResource centres, NHS Trusts and staff for their contribution. We thank the National Institute for Health Research, NHS Blood and Transplant, and Health Data Research UK as part of the Digital Innovation Hub Programme. This study presents independent research funded by the NIHR Biomedical Research Centre at South London and Maudsley NHS Foundation Trust and King’s College London. Further information can be found at http://brc.slam.nhs.uk/about/core-facilities/bioresource. The views expressed are those of the authors and not necessarily those of the NHS, the NIHR, the HSC R&D Division, King’s College London, or the Department of Health and Social Care.

The PROTECT study was funded/supported by the National Institute of Health and Care Research Exeter Biomedical Research Centre. PROTECT genetic data was funded in part by the University of Exeter through the MRC Proximity to Discovery: Industry Engagement Fund (External Collaboration, Innovation and Entrepreneurism: Translational Medicine in Exeter 2 (EXCITEME2) ref. MC_PC_17189). Genotyping was performed at deCODE Genetics. The views expressed are those of the author(s) and not necessarily those of the NIHR or the Dept of Health and Social Care.

As data used in this study was obtained from the ADNI database, the ADNI investigators contributed to the conception of the sample and acquisition of this data, although were not participants in other parts of this study, such as conceptualisation, data analysis or writing. A full acknowledgment list of ADNI investigators can be found at: https://adni.loni.usc.edu/wp-content/uploads/how_to_apply/ADNI_Acknowledgement_List.pdf

Similarly, data was obtained from the GERAD1 Consortium and GERAD1 investigators contributed to the conception of the cohort and acquisition of this data, but did not participate in the conceptualisation, analysis or writing of this study. A full list of GERAD1 collaborators can be found in Supplementary Material 3.

We also thank and acknowledge the contribution and use of the CREATE high-performance computing cluster at King’s College London (King’s College London. (2022). King’s Computational Research, Engineering and Technology Environment (CREATE). Retrieved May 23, 2023, from https://doi.org/10.18742/rnvf-m076).

The analysis flowchart in Figure 1. was created and licenced with BioRender (www.biorender.com)

## 6. Conflicts of Interest

CML is on the Scientific Advisory Board of Myriad Neuroscience and has received honoraria for consultancy from UCB.

## 7. Data availability

This study made use of publicly available analysis software:

MungeSumstats (https://neurogenomics.github.io/MungeSumstats/articles/MungeSumstats.html)

REGENIE (https://rgcgithub.github.io/regenie/)

MTAG (https://github.com/JonJala/mtag)

METAL (https://genome.sph.umich.edu/wiki/METAL_Documentation)

FUMA GWAS (https://fuma.ctglab.nl)

LDSC (https://github.com/bulik/ldsc)

HDL (https://github.com/zhenin/HDL)

LAVA (https://github.com/josefin-werme/LAVA)

susieR (https://stephenslab.github.io/susieR/index.html)

coloc (https://chr1swallace.github.io/coloc/)

COLOC-reporter (https://github.com/ThomasPSpargo/COLOC-reporter)

CAUSE (https://jean997.github.io/cause/index.html)

TwoSampleMR (https://mrcieu.github.io/TwoSampleMR/)

MegaPRS (https://dougspeed.com/megaprs/)

## Notes

### Funding Statement

See funding and acknowledgement statement in the main body of the manuscript

### Author Declarations

Ethics committee/IRB of King's College London gave ethical approval for this work Ethical approval for the UK Biobank study has been granted by the National Information Governance Board for Health and Social Care and the NHS North West Multicentre Research Ethics Committee (11/NW/0382). Data access permission has been granted under UK Biobank application 18177. Written informed consent was obtained from all participants by the UK Biobank. The GLAD Study was approved by the London - Fulham Research Ethics Committee on 21st August 2018 (REC reference: 18/LO/1218) following a full review by the committee. The NIHR BioResource has been approved as a Research Tissue Bank by the East of England - Cambridge Central Committee (REC reference: 17/EE/0025). The PROTECT study received ethical approval from the UK London Bridge National Research Ethics Committee (Ref: 13/LO/1578) Data were obtained from ADNI, AddNeuroMed and GERAD1 following formal request to each consortia. Permission was granted for the use of data.

### Summary of Updates

Author added; author affiliations updated; supplementary figures added

## References

1. Byers, A.L., and Yaffe, K. (2011). Depression and risk of developing dementia. Nat. Rev. Neurol. 7, 323–331.

2. Livingston, G., Huntley, J., Sommerlad, A., Ames, D., Ballard, C., Banerjee, S., Brayne, C., Burns, A., Cohen-Mansfield, J., Cooper, C., et al. (2020). Dementia prevention, intervention, and care: 2020 report of the Lancet Commission. Lancet 396, 413–446.

3. Yu, O.-C., Jung, B., Go, H., Park, M., and Ha, I.-H. (2020). Association between dementia and depression: a retrospective study using the Korean National Health Insurance Service-National Sample Cohort database. BMJ Open 10, e034924.

4. Yang, W., Li, X., Pan, K.-Y., Yang, R., Song, R., Qi, X., Pedersen, N.L., and Xu, W. (2021). Association of life-course depression with the risk of dementia in late life: A nationwide twin study. Alzheimers Dement. 17, 1383–1390.

5. Richmond-Rakerd, L.S., D’Souza, S., Milne, B.J., Caspi, A., and Moffitt, T.E. (2022). Longitudinal Associations of Mental Disorders With Dementia: 30-Year Analysis of 1.7 Million New Zealand Citizens. JAMA Psychiatry 79, 333–340.

6. Yang, L., Deng, Y.-T., Leng, Y., Ou, Y.-N., Li, Y.-Z., Chen, S.-D., He, X.-Y., Wu, B.-S., Huang, S.-Y., Zhang, Y.-R., et al. (2022). Depression, depression treatments, and risk of incident dementia: A prospective cohort study of 354,313 participants. Biol. Psychiatry.

7. GBD 2016 Dementia Collaborators (2019). Global, regional, and national burden of Alzheimer’s disease and other dementias, 1990-2016: a systematic analysis for the Global Burden of Disease Study 2016. Lancet Neurol. 18, 88–106.

8. Dafsari, F.S., and Jessen, F. (2020). Depression-an underrecognized target for prevention of dementia in Alzheimer’s disease. Transl. Psychiatry 10, 160.

9. Corfield, E.C., Yang, Y., Martin, N.G., and Nyholt, D.R. (2017). A continuum of genetic liability for minor and major depression. Transl. Psychiatry 7, e1131.

10. Gatz, M., Reynolds, C.A., Fratiglioni, L., Johansson, B., Mortimer, J.A., Berg, S., Fiske, A., and Pedersen, N.L. (2006). Role of genes and environments for explaining Alzheimer disease. Arch. Gen. Psychiatry 63, 168–174.

11. Wray, N.R., Ripke, S., Mattheisen, M., Trzaskowski, M., Byrne, E.M., Abdellaoui, A., Adams, M.J., Agerbo, E., Air, T.M., Andlauer, T.M.F., et al. (2018). Genome-wide association analyses identify 44 risk variants and refine the genetic architecture of major depression. Nat. Genet. 50, 668–681.

12. Howard, D.M., Adams, M.J., Clarke, T.-K., Hafferty, J.D., Gibson, J., Shirali, M., Coleman, J.R.I., Hagenaars, S.P., Ward, J., Wigmore, E.M., et al. (2019). Genome-wide meta-analysis of depression identifies 102 independent variants and highlights the importance of the prefrontal brain regions. Nat. Neurosci. 22, 343–352.

13. Levey, D.F., Stein, M.B., Wendt, F.R., Pathak, G.A., Zhou, H., Aslan, M., Quaden, R., Harrington, K.M., Nuñez, Y.Z., Overstreet, C., et al. (2021). Bi-ancestral depression GWAS in the Million Veteran Program and meta-analysis in >1.2 million individuals highlight new therapeutic directions. Nat. Neurosci. 24, 954–963.

14. Kunkle, B.W., Grenier-Boley, B., Sims, R., Bis, J.C., Damotte, V., Naj, A.C., Boland, A., Vronskaya, M., van der Lee, S.J., Amlie-Wolf, A., et al. (2019). Genetic meta-analysis of diagnosed Alzheimer’s disease identifies new risk loci and implicates Aβ, tau, immunity and lipid processing. Nat. Genet. 51, 414–430.

15. Jansen, I.E., Savage, J.E., Watanabe, K., Bryois, J., Williams, D.M., Steinberg, S., Sealock, J., Karlsson, I.K., Hägg, S., Athanasiu, L., et al. (2019). Genome-wide meta-analysis identifies new loci and functional pathways influencing Alzheimer’s disease risk. Nat. Genet. 51, 404–413.

16. Wightman, D.P., Jansen, I.E., Savage, J.E., Shadrin, A.A., Bahrami, S., Holland, D., Rongve, A., Børte, S., Winsvold, B.S., Drange, O.K., et al. (2021). A genome-wide association study with 1,126,563 individuals identifies new risk loci for Alzheimer’s disease. Nat. Genet. 53, 1276–1282.

17. Bellenguez, C., Küçükali, F., Jansen, I.E., Kleineidam, L., Moreno-Grau, S., Amin, N., Naj, A.C., Campos-Martin, R., Grenier-Boley, B., Andrade, V., et al. (2022). New insights into the genetic etiology of Alzheimer’s disease and related dementias. Nat. Genet. 54, 412– 436.

18. Gibson, J., Russ, T.C., Adams, M.J., Clarke, T.K., Howard, D.M., Hall, L.S., Fernandez-Pujals, A.M., Wigmore, E.M., Hayward, C., Davies, G., et al. (2017). Assessing the presence of shared genetic architecture between Alzheimer’s disease and major depressive disorder using genome-wide association data. Transl. Psychiatry 7, e1094.

19. Andrews, S.J., Fulton-Howard, B., O’Reilly, P., Marcora, E., Goate, A.M., and collaborators of the Alzheimer’s Disease Genetics Consortium (2021). Causal associations between modifiable risk factors and the alzheimer’s phenome. Ann. Neurol. 89, 54–65.

20. Santos, F.C.D., Mendes-Silva, A.P., Nikolova, Y.C., Sibille, E., and Diniz, B.S. (2021). Genetic overlap between major depression, bipolar disorder and Alzheimer’s Disease. MedRxiv.

21. Monereo-Sánchez, J., Schram, M.T., Frei, O., O’Connell, K., Shadrin, A.A., Smeland, O.B., Westlye, L.T., Andreassen, O.A., Kaufmann, T., Linden, D.E.J., et al. (2021). Genetic overlap between alzheimer’s disease and depression mapped onto the brain. Front. Neurosci. 15, 653130.

22. Harerimana, N.V., Liu, Y., Gerasimov, E.S., Duong, D., Beach, T.G., Reiman, E.M., Schneider, J.A., Boyle, P., Lori, A., Bennett, D.A., et al. (2022). Genetic evidence supporting a causal role of depression in alzheimer’s disease. Biol. Psychiatry 92, 25–33.

23. American Psychiatric Association (2013). Depressive Disorders. In Diagnostic and Statistical Manual of Mental Disorders, (American Psychiatric Association), p.

24. Fried, E.I., and Nesse, R.M. (2015). Depression is not a consistent syndrome: An investigation of unique symptom patterns in the STAR*D study. J. Affect. Disord. 172, 96– 102.

25. Zimmerman, M., Ellison, W., Young, D., Chelminski, I., and Dalrymple, K. (2015). How many different ways do patients meet the diagnostic criteria for major depressive disorder? Compr. Psychiatry 56, 29–34.

26. Park, S.-C., Kim, J.-M., Jun, T.-Y., Lee, M.-S., Kim, J.-B., Yim, H.-W., and Park, Y.C. (2017). How many different symptom combinations fulfil the diagnostic criteria for major depressive disorder? Results from the CRESCEND study. Nord J Psychiatry 71, 217–222.

27. Cai, N., Choi, K.W., and Fried, E.I. (2020). Reviewing the genetics of heterogeneity in depression: operationalizations, manifestations and etiologies. Hum. Mol. Genet. 29, R10– R18.

28. Nguyen, T.-D., Harder, A., Xiong, Y., Kowalec, K., Hägg, S., Cai, N., Kuja-Halkola, R., Dalman, C., Sullivan, P.F., and Lu, Y. (2022). Genetic heterogeneity and subtypes of major depression. Mol. Psychiatry 27, 1667–1675.

29. Thorp, J.G., Marees, A.T., Ong, J.-S., An, J., MacGregor, S., and Derks, E.M. (2020). Genetic heterogeneity in self-reported depressive symptoms identified through genetic analyses of the PHQ-9. Psychol. Med. 50, 2385–2396.

30. So, Y., Kim, K.W., Park, J.H., Lee, J.J., Lee, S.B., and TaeHui Kim (2012). P3-396: Anhedonia is associated with the risk of Alzheimer’s disease in elders with mild cognitive impairment: Results from the Korean Longitudinal Study on Health and Aging (KLOSHA). Alzheimers Dement. 8, P594–P594.

31. Lee, J.R., Suh, S.W., Han, J.W., Byun, S., Kwon, S.J., Lee, K.H., Kwak, K.P., Kim, B.J., Kim, S.G., Kim, J.L., et al. (2019). Anhedonia and Dysphoria Are Differentially Associated with the Risk of Dementia in the Cognitively Normal Elderly Individuals: A Prospective Cohort Study. Psychiatry Investig 16, 575–580.

32. Vaquero-Puyuelo, D., De-la-Cámara, C., Olaya, B., Gracia-García, P., Lobo, A., López-Antón, R., and Santabárbara, J. (2021). Anhedonia as a Potential Risk Factor of Alzheimer’s Disease in a Community-Dwelling Elderly Sample: Results from the ZARADEMP Project. Int. J. Environ. Res. Public Health 18,.

33. Ikeda, M., Brown, J., Holland, A.J., Fukuhara, R., and Hodges, J.R. (2002). Changes in appetite, food preference, and eating habits in frontotemporal dementia and Alzheimer’s disease. J. Neurol. Neurosurg. Psychiatry 73, 371–376.

34. Bailon, O., Roussel, M., Boucart, M., Krystkowiak, P., and Godefroy, O. (2010). Psychomotor slowing in mild cognitive impairment, Alzheimer’s disease and lewy body dementia: mechanisms and diagnostic value. Dement. Geriatr. Cogn. Disord. 29, 388–396.

35. Koren, T., Fisher, E., Webster, L., Livingston, G., and Rapaport, P. (2022). Prevalence of sleep disturbances in people with dementia living in the community: A systematic review and meta-analysis. Ageing Res Rev 101782.

36. Escott-Price, V., and Hardy, J. (2022). Genome-wide association studies for Alzheimer’s disease: bigger is not always better. Brain Commun. 4, fcac125.

37. Marioni, R.E., Harris, S.E., Zhang, Q., McRae, A.F., Hagenaars, S.P., Hill, W.D., Davies, G., Ritchie, C.W., Gale, C.R., Starr, J.M., et al. (2018). GWAS on family history of Alzheimer’s disease. Transl. Psychiatry 8, 99.

38. Liu, H., Hu, Y., Zhang, Y., Zhang, H., Gao, S., Wang, L., Wang, T., Han, Z., Sun, B.-L., and Liu, G. (2022). Mendelian randomization highlights significant difference and genetic heterogeneity in clinically diagnosed Alzheimer’s disease GWAS and self-report proxy phenotype GWAX. Alzheimers Res. Ther. 14, 17.

39. Desai, R., John, A., Saunders, R., Marchant, N.L., Buckman, J.E.J., Charlesworth, G., Zuber, V., and Stott, J. (2023). Examining the Lancet Commission risk factors for dementia using Mendelian randomisation. BMJ Ment Health 26,.

40. European Alzheimer’s & Dementia Biobank Mendelian Randomization (EADB-MR) Collaboration, Luo, J., Thomassen, J.Q., Bellenguez, C., Grenier-Boley, B., de Rojas, I., Castillo, A., Parveen, K., Küçükali, F., Nicolas, A., et al. (2023). Genetic associations between modifiable risk factors and alzheimer disease. JAMA Netw. Open 6, e2313734.

41. Davies, M.R., Kalsi, G., Armour, C., Jones, I.R., McIntosh, A.M., Smith, D.J., Walters, J.T.R., Bradley, J.R., Kingston, N., Ashford, S., et al. (2019). The Genetic Links to Anxiety and Depression (GLAD) Study: Online recruitment into the largest recontactable study of depression and anxiety. Behav. Res. Ther. 123, 103503.

42. Creese, B., Arathimos, R., Brooker, H., Aarsland, D., Corbett, A., Lewis, C., Ballard, C., and Ismail, Z. (2021). Genetic risk for Alzheimer’s disease, cognition, and mild behavioral impairment in healthy older adults. Alzheimers Dement (Amst) 13, e12164.

43. Arathimos, R., Fabbri, C., Vassos, E., Davis, K.A.S., Pain, O., Gillett, A., Coleman, J.R.I., Hanscombe, K., Hagenaars, S., Jermy, B., et al. (2022). Latent subtypes of manic and/or irritable episode symptoms in two population-based cohorts. Br. J. Psychiatry 221, 722–731.

44. Bycroft, C., Freeman, C., Petkova, D., Band, G., Elliott, L.T., Sharp, K., Motyer, A., Vukcevic, D., Delaneau, O., O’Connell, J., et al. (2018). The UK Biobank resource with deep phenotyping and genomic data. Nature 562, 203–209.

45. Wightman, D.P., Savage, J.E., Tissink, E., Romero, C., Jansen, I.E., and Posthuma, D. (2023). The genetic overlap between Alzheimer’s disease, amyotrophic lateral sclerosis, Lewy body dementia, and Parkinson’s disease. Neurobiol. Aging.

46. Kroenke, K., Spitzer, R.L., and Williams, J.B. (2001). The PHQ-9: validity of a brief depression severity measure. J. Gen. Intern. Med. 16, 606–613.

47. Sun, Y., Fu, Z., Bo, Q., Mao, Z., Ma, X., and Wang, C. (2020). The reliability and validity of PHQ-9 in patients with major depressive disorder in psychiatric hospital. BMC Psychiatry 20, 474.

48. Mbatchou, J., Barnard, L., Backman, J., Marcketta, A., Kosmicki, J.A., Ziyatdinov, A., Benner, C., O’Dushlaine, C., Barber, M., Boutkov, B., et al. (2021). Computationally efficient whole-genome regression for quantitative and binary traits. Nat. Genet. 53, 1097– 1103.

49. Chang, C.C., Chow, C.C., Tellier, L.C., Vattikuti, S., Purcell, S.M., and Lee, J.J. (2015). Second-generation PLINK: rising to the challenge of larger and richer datasets. Gigascience 4, 7.

50. Privé, F., Luu, K., Blum, M.G.B., McGrath, J.J., and Vilhjálmsson, B.J. (2020). Efficient toolkit implementing best practices for principal component analysis of population genetic data. Bioinformatics 36, 4449–4457.

51. McCarthy, S., Das, S., Kretzschmar, W., Delaneau, O., Wood, A.R., Teumer, A., Kang, H.M., Fuchsberger, C., Danecek, P., Sharp, K., et al. (2016). A reference panel of 64,976 haplotypes for genotype imputation. Nat. Genet. 48, 1279–1283.

52. Chou, W.-C., Zheng, H.-F., Cheng, C.-H., Yan, H., Wang, L., Han, F., Richards, J.B., Karasik, D., Kiel, D.P., and Hsu, Y.-H. (2016). A combined reference panel from the 1000 Genomes and UK10K projects improved rare variant imputation in European and Chinese samples. Sci. Rep. 6, 39313.

53. Taliun, D., Harris, D.N., Kessler, M.D., Carlson, J., Szpiech, Z.A., Torres, R., Taliun, S.A.G., Corvelo, A., Gogarten, S.M., Kang, H.M., et al. (2021). Sequencing of 53,831 diverse genomes from the NHLBI TOPMed Program. Nature 590, 290–299.

54. 1000 Genomes Project Consortium, Auton, A., Brooks, L.D., Durbin, R.M., Garrison, E.P., Kang, H.M., Korbel, J.O., Marchini, J.L., McCarthy, S., McVean, G.A., et al. (2015). A global reference for human genetic variation. Nature 526, 68–74.

55. Murphy, A.E., Schilder, B.M., and Skene, N.G. (2021). MungeSumstats: A Bioconductor package for the standardisation and quality control of many GWAS summary statistics. Bioinformatics.

56. Bulik-Sullivan, B., Loh, P.-R., Finucane, H.K., Ripke, S., Yang, J., Schizophrenia Working Group of the Psychiatric Genomics Consortium, Patterson, N., Daly, M.J., Price, A.L., and Neale, B.M. (2015). LD Score regression distinguishes confounding from polygenicity in genome-wide association studies. Nat. Genet. 47, 291–295.

57. Bulik-Sullivan, B., Finucane, H.K., Anttila, V., Gusev, A., Day, F.R., Loh, P.-R., ReproGen Consortium, Psychiatric Genomics Consortium, Genetic Consortium for Anorexia Nervosa of the Wellcome Trust Case Control Consortium 3, Duncan, L., et al. (2015). An atlas of genetic correlations across human diseases and traits. Nat. Genet. 47, 1236–1241.

58. Willer, C.J., Li, Y., and Abecasis, G.R. (2010). METAL: fast and efficient meta-analysis of genomewide association scans. Bioinformatics 26, 2190–2191.

59. Turley, P., Walters, R.K., Maghzian, O., Okbay, A., Lee, J.J., Fontana, M.A., Nguyen-Viet, T.A., Wedow, R., Zacher, M., Furlotte, N.A., et al. (2018). Multi-trait analysis of genome-wide association summary statistics using MTAG. Nat. Genet. 50, 229–237.

60. Watanabe, K., Taskesen, E., van Bochoven, A., and Posthuma, D. (2017). Functional mapping and annotation of genetic associations with FUMA. Nat. Commun. 8, 1826.

61. Jaffe, A.E., Straub, R.E., Shin, J.H., Tao, R., Gao, Y., Collado-Torres, L., Kam-Thong, T., Xi, H.S., Quan, J., Chen, Q., et al. (2018). Developmental and genetic regulation of the human cortex transcriptome illuminate schizophrenia pathogenesis. Nat. Neurosci. 21, 1117– 1125.

62. Wang, D., Liu, S., Warrell, J., Won, H., Shi, X., Navarro, F.C.P., Clarke, D., Gu, M., Emani, P., Yang, Y.T., et al. (2018). Comprehensive functional genomic resource and integrative model for the human brain. Science 362,.

63. Fromer, M., Roussos, P., Sieberts, S.K., Johnson, J.S., Kavanagh, D.H., Perumal, T.M., Ruderfer, D.M., Oh, E.C., Topol, A., Shah, H.R., et al. (2016). Gene expression elucidates functional impact of polygenic risk for schizophrenia. Nat. Neurosci. 19, 1442–1453.

64. Ramasamy, A., Trabzuni, D., Guelfi, S., Varghese, V., Smith, C., Walker, R., De, T., UK Brain Expression Consortium, North American Brain Expression Consortium, Coin, L., et al. (2014). Genetic variability in the regulation of gene expression in ten regions of the human brain. Nat. Neurosci. 17, 1418–1428.

65. Westra, H.-J., Peters, M.J., Esko, T., Yaghootkar, H., Schurmann, C., Kettunen, J., Christiansen, M.W., Fairfax, B.P., Schramm, K., Powell, J.E., et al. (2013). Systematic identification of trans eQTLs as putative drivers of known disease associations. Nat. Genet. 45, 1238–1243.

66. Zhernakova, D.V., Deelen, P., Vermaat, M., van Iterson, M., van Galen, M., Arindrarto, W., van ’t Hof, P., Mei, H., van Dijk, F., Westra, H.-J., et al. (2017). Identification of context-dependent expression quantitative trait loci in whole blood. Nat. Genet. 49, 139–145.

67. Võsa, U., Claringbould, A., Westra, H.-J., Bonder, M.J., Deelen, P., Zeng, B., Kirsten, H., Saha, A., Kreuzhuber, R., Kasela, S., et al. (2018). Unraveling the polygenic architecture of complex traits using blood eQTL meta-analysis. BioRxiv.

68. Buil, A., Brown, A.A., Lappalainen, T., Viñuela, A., Davies, M.N., Zheng, H.-F., Richards, J.B., Glass, D., Small, K.S., Durbin, R., et al. (2015). Gene-gene and gene-environment interactions detected by transcriptome sequence analysis in twins. Nat. Genet. 47, 88–91.

69. Ng, B., White, C.C., Klein, H.-U., Sieberts, S.K., McCabe, C., Patrick, E., Xu, J., Yu, L., Gaiteri, C., Bennett, D.A., et al. (2017). An xQTL map integrates the genetic architecture of the human brain’s transcriptome and epigenome. Nat. Neurosci. 20, 1418–1426.

70. van Rheenen, W., Peyrot, W.J., Schork, A.J., Lee, S.H., and Wray, N.R. (2019). Genetic correlations of polygenic disease traits: from theory to practice. Nat. Rev. Genet. 20, 567– 581.

71. Ning, Z., Pawitan, Y., and Shen, X. (2020). High-definition likelihood inference of genetic correlations across human complex traits. Nat. Genet. 52, 859–864.

72. Werme, J., van der Sluis, S., Posthuma, D., and de Leeuw, C.A. (2022). An integrated framework for local genetic correlation analysis. Nat. Genet. 54, 274–282.

73. Spargo, T.P., Gilchrist, L., Hunt, G.P., Dobson, R.J., Proitsi, P.P., Al-Chalabi, A., Pain, O., and Iacoangeli, A. (2023). Statistical examination of shared loci in neuropsychiatric diseases using genome-wide association study summary statistics. MedRxiv.

74. Zou, Y., Carbonetto, P., Wang, G., and Stephens, M. (2022). Fine-mapping from summary data with the “Sum of Single Effects” model. PLoS Genet. 18, e1010299.

75. Wallace, C. (2021). A more accurate method for colocalisation analysis allowing for multiple causal variants. PLoS Genet. 17, e1009440.

76. Giambartolomei, C., Vukcevic, D., Schadt, E.E., Franke, L., Hingorani, A.D., Wallace, C., and Plagnol, V. (2014). Bayesian test for colocalisation between pairs of genetic association studies using summary statistics. PLoS Genet. 10, e1004383.

77. Sanderson, E., Glymour, M.M., Holmes, M.V., Kang, H., Morrison, J., Munafò, M.R., Palmer, T., Schooling, C.M., Wallace, C., Zhao, Q., et al. (2022). Mendelian randomization. Nat. Rev. Methods Primers 2, 6.

78. Burgess, S., Davies, N.M., and Thompson, S.G. (2016). Bias due to participant overlap in two-sample Mendelian randomization. Genet. Epidemiol. 40, 597–608.

79. Morrison, J., Knoblauch, N., Marcus, J.H., Stephens, M., and He, X. (2020). Mendelian randomization accounting for correlated and uncorrelated pleiotropic effects using genome-wide summary statistics. Nat. Genet. 52, 740–747.

80. Hemani, G., Bowden, J., and Davey Smith, G. (2018). Evaluating the potential role of pleiotropy in Mendelian randomization studies. Hum. Mol. Genet. 27, R195–R208.

81. Slob, E.A.W., and Burgess, S. (2020). A comparison of robust Mendelian randomization methods using summary data. Genet. Epidemiol. 44, 313–329.

82. Bowden, J., Davey Smith, G., and Burgess, S. (2015). Mendelian randomization with invalid instruments: effect estimation and bias detection through Egger regression. Int. J. Epidemiol. 44, 512–525.

83. Bowden, J., Davey Smith, G., Haycock, P.C., and Burgess, S. (2016). Consistent Estimation in Mendelian Randomization with Some Invalid Instruments Using a Weighted Median Estimator. Genet. Epidemiol. 40, 304–314.

84. Hartwig, F.P., Davey Smith, G., and Bowden, J. (2017). Robust inference in summary data Mendelian randomization via the zero modal pleiotropy assumption. Int. J. Epidemiol. 46, 1985–1998.

85. Verbanck, M., Chen, C.-Y., Neale, B., and Do, R. (2018). Detection of widespread horizontal pleiotropy in causal relationships inferred from Mendelian randomization between complex traits and diseases. Nat. Genet. 50, 693–698.

86. Burgess, S., and Thompson, S.G. (2017). Interpreting findings from Mendelian randomization using the MR-Egger method. Eur J Epidemiol 32, 377–389.

87. Burgess, S., Bowden, J., Fall, T., Ingelsson, E., and Thompson, S.G. (2017). Sensitivity Analyses for Robust Causal Inference from Mendelian Randomization Analyses with Multiple Genetic Variants. Epidemiology 28, 30–42.

88. Lawlor, D.A., Harbord, R.M., Sterne, J.A.C., Timpson, N., and Davey Smith, G. (2008). Mendelian randomization: using genes as instruments for making causal inferences in epidemiology. Stat. Med. 27, 1133–1163.

89. Bowden, J., Del Greco M, F., Minelli, C., Davey Smith, G., Sheehan, N.A., and Thompson, J.R. (2016). Assessing the suitability of summary data for two-sample Mendelian randomization analyses using MR-Egger regression: the role of the I2 statistic. Int. J. Epidemiol. 45, 1961–1974.

90. Mahley, R.W. (2016). Apolipoprotein E: from cardiovascular disease to neurodegenerative disorders. J. Mol. Med. 94, 739–746.

91. Liu, S., Liu, J., Weng, R., Gu, X., and Zhong, Z. (2019). Apolipoprotein E gene polymorphism and the risk of cardiovascular disease and type 2 diabetes. BMC Cardiovasc. Disord. 19, 213.

92. Xuan, L., Zhao, Z., Jia, X., Hou, Y., Wang, T., Li, M., Lu, J., Xu, Y., Chen, Y., Qi, L., et al. (2018). Type 2 diabetes is causally associated with depression: a Mendelian randomization analysis. Front Med 12, 678–687.

93. Lu, Y., Wang, Z., Georgakis, M.K., Lin, H., and Zheng, L. (2021). Genetic liability to depression and risk of coronary artery disease, myocardial infarction, and other cardiovascular outcomes. J. Am. Heart Assoc. 10, e017986.

94. Lord, J., Jermy, B., Green, R., Wong, A., Xu, J., Legido-Quigley, C., Dobson, R., Richards, M., and Proitsi, P. (2021). Mendelian randomization identifies blood metabolites previously linked to midlife cognition as causal candidates in Alzheimer’s disease. Proc. Natl. Acad. Sci. USA 118,.

95. Choi, S.W., Mak, T.S.-H., and O’Reilly, P.F. (2020). Tutorial: a guide to performing polygenic risk score analyses. Nat. Protoc. 15, 2759–2772.

96. Zhang, Q., Privé, F., Vilhjálmsson, B., and Speed, D. (2021). Improved genetic prediction of complex traits from individual-level data or summary statistics. Nat. Commun. 12, 4192.

97. Lovestone, S., Francis, P., Kloszewska, I., Mecocci, P., Simmons, A., Soininen, H., Spenger, C., Tsolaki, M., Vellas, B., Wahlund, L.-O., et al. (2009). AddNeuroMed--the European collaboration for the discovery of novel biomarkers for Alzheimer’s disease. Ann. N. Y. Acad. Sci. 1180, 36–46.

98. Lord, J., Green, R., Choi, S.W., Hübel, C., Aarsland, D., Velayudhan, L., Sham, P., Legido-Quigley, C., Richards, M., Dobson, R., et al. (2022). Disentangling independent and mediated causal relationships between blood metabolites, cognitive factors, and alzheimer’s disease. Biological Psychiatry Global Open Science 2, 167–179.

99. Yang, J., Weedon, M.N., Purcell, S., Lettre, G., Estrada, K., Willer, C.J., Smith, A.V., Ingelsson, E., O’Connell, J.R., Mangino, M., et al. (2011). Genomic inflation factors under polygenic inheritance. Eur. J. Hum. Genet. 19, 807–812.

100. Voskobiynyk, Y., Roth, J.R., Cochran, J.N., Rush, T., Carullo, N.V., Mesina, J.S., Waqas, M., Vollmer, R.M., Day, J.J., McMahon, L.L., et al. (2020). Alzheimer’s disease risk gene BIN1 induces Tau-dependent network hyperexcitability. Elife 9,.

101. Lambert, E., Saha, O., Soares Landeira, B., Melo de Farias, A.R., Hermant, X., Carrier, A., Pelletier, A., Gadaut, J., Davoine, L., Dupont, C., et al. (2022). The Alzheimer susceptibility gene BIN1 induces isoform-dependent neurotoxicity through early endosome defects. Acta Neuropathol. Commun. 10, 4.

102. Pruim, R.J., Welch, R.P., Sanna, S., Teslovich, T.M., Chines, P.S., Gliedt, T.P., Boehnke, M., Abecasis, G.R., and Willer, C.J. (2010). LocusZoom: regional visualization of genome-wide association scan results. Bioinformatics 26, 2336–2337.

103. Dashti, H.S., Daghlas, I., Lane, J.M., Huang, Y., Udler, M.S., Wang, H., Ollila, H.M., Jones, S.E., Kim, J., Wood, A.R., et al. (2021). Genetic determinants of daytime napping and effects on cardiometabolic health. Nat. Commun. 12, 900.

104. Jansen, P.R., Watanabe, K., Stringer, S., Skene, N., Bryois, J., Hammerschlag, A.R., de Leeuw, C.A., Benjamins, J.S., Muñoz-Manchado, A.B., Nagel, M., et al. (2019). Genome-wide analysis of insomnia in 1,331,010 individuals identifies new risk loci and functional pathways. Nat. Genet. 51, 394–403.

105. Hammerschlag, A.R., Stringer, S., de Leeuw, C.A., Sniekers, S., Taskesen, E., Watanabe, K., Blanken, T.F., Dekker, K., Te Lindert, B.H.W., Wassing, R., et al. (2017). Genome-wide association analysis of insomnia complaints identifies risk genes and genetic overlap with psychiatric and metabolic traits. Nat. Genet. 49, 1584–1592.

106. Loos, R.J.F., and Yeo, G.S.H. (2022). The genetics of obesity: from discovery to biology. Nat. Rev. Genet. 23, 120–133.

107. Zarza-Rebollo, J.A., Molina, E., and Rivera, M. (2021). The role of the FTO gene in the relationship between depression and obesity. A systematic review. Neurosci. Biobehav. Rev. 127, 630–637.

108. González, L.M., García-Herráiz, A., Mota-Zamorano, S., Flores, I., Albuquerque, D., and Gervasini, G. (2021). Variants in the Obesity-Linked FTO gene locus modulates psychopathological features of patients with Anorexia Nervosa. Gene 783, 145572.

109. Li, Y., Dang, X., Chen, R., Wang, J., Li, S., Mitchell, B.L., Yao, Y.-G., Li, M., Li, T., Zhang, Z., et al. (2023). Cross-ancestry genome-wide association study and systems-level integrative analyses implicate new risk genes and therapeutic targets for depression. MedRxiv.

110. Dall’Aglio, L., Lewis, C.M., and Pain, O. (2021). Delineating the genetic component of gene expression in major depression. Biol. Psychiatry 89, 627–636.

111. Fabbri, C., Pain, O., Hagenaars, S.P., Lewis, C.M., and Serretti, A. (2021). Transcriptome-wide association study of treatment-resistant depression and depression subtypes for drug repurposing. Neuropsychopharmacology 46, 1821–1829.

112. Koskinen, M.-K., and Hovatta, I. (2023). Genetic insights into the neurobiology of anxiety. Trends Neurosci. 46, 318–331.

113. Nagel, M., Jansen, P.R., Stringer, S., Watanabe, K., de Leeuw, C.A., Bryois, J., Savage, J.E., Hammerschlag, A.R., Skene, N.G., Muñoz-Manchado, A.B., et al. (2018). Meta-analysis of genome-wide association studies for neuroticism in 449,484 individuals identifies novel genetic loci and pathways. Nat. Genet. 50, 920–927.

114. Mallard, T.T., Linnér, R.K., Grotzinger, A.D., Sanchez-Roige, S., Seidlitz, J., Okbay, A., de Vlaming, R., Meddens, S.F.W., Bipolar Disorder Working Group of the Psychiatric Genomics Consortium, Palmer, A.A., et al. (2022). Multivariate GWAS of psychiatric disorders and their cardinal symptoms reveal two dimensions of cross-cutting genetic liabilities. Cell Genomics 2,.

115. Lüningschrör, P., Werner, G., Stroobants, S., Kakuta, S., Dombert, B., Sinske, D., Wanner, R., Lüllmann-Rauch, R., Wefers, B., Wurst, W., et al. (2020). The FTLD risk factor TMEM106B regulates the transport of lysosomes at the axon initial segment of motoneurons. Cell Rep. 30, 3506–3519.e6.

116. Van Deerlin, V.M., Sleiman, P.M.A., Martinez-Lage, M., Chen-Plotkin, A., Wang, L.-S., Graff-Radford, N.R., Dickson, D.W., Rademakers, R., Boeve, B.F., Grossman, M., et al. (2010). Common variants at 7p21 are associated with frontotemporal lobar degeneration with TDP-43 inclusions. Nat. Genet. 42, 234–239.

117. Nicholson, A.M., and Rademakers, R. (2016). What we know about TMEM106B in neurodegeneration. Acta Neuropathol. 132, 639–651.

118. Feng, T., Lacrampe, A., and Hu, F. (2021). Physiological and pathological functions of TMEM106B: a gene associated with brain aging and multiple brain disorders. Acta Neuropathol. 141, 327–339.

119. Perneel, J., and Rademakers, R. (2022). Identification of TMEM106B amyloid fibrils provides an updated view of TMEM106B biology in health and disease. Acta Neuropathol. 144, 807–819.

120. Mao, F., Robinson, J.L., Unger, T., Posavi, M., Amado, D.A., Elman, L., Grossman, M., Wolk, D.A., Lee, E.B., Van Deerlin, V.M., et al. (2021). TMEM106B modifies TDP-43 pathology in human ALS brain and cell-based models of TDP-43 proteinopathy. Acta Neuropathol. 142, 629–642.

121. Vass, R., Ashbridge, E., Geser, F., Hu, W.T., Grossman, M., Clay-Falcone, D., Elman, L., McCluskey, L., Lee, V.M.Y., Van Deerlin, V.M., et al. (2011). Risk genotypes at TMEM106B are associated with cognitive impairment in amyotrophic lateral sclerosis. Acta Neuropathol. 121, 373–380.

122. Shafit-Zagardo, B., Sidoli, S., Goldman, J.E., DuBois, J.C., Corboy, J.R., Strittmatter, S.M., Guzik, H., Graff, S., and Nagra, R.M. (2022). TMEM106B is increased in Multiple Sclerosis plaques, and deletion causes accumulation of lipid after demyelination. BioRxiv.

123. Hong, S., Dobricic, V., Ohlei, O., Bos, I., Vos, S.J.B., Prokopenko, D., Tijms, B.M., Andreasson, U., Blennow, K., Vandenberghe, R., et al. (2021). TMEM106B and CPOX are genetic determinants of cerebrospinal fluid Alzheimer’s disease biomarker levels. Alzheimers Dement. 17, 1628–1640.

124. Dhiman, K., Gupta, V.B., Villemagne, V.L., Eratne, D., Graham, P.L., Fowler, C., Bourgeat, P., Li, Q.-X., Collins, S., Bush, A.I., et al. (2020). Cerebrospinal fluid neurofilament light concentration predicts brain atrophy and cognition in Alzheimer’s disease. Alzheimers Dement (Amst) 12, e12005.

125. Chen, M.-H., Liu, Y.-L., Kuo, H.-W., Tsai, S.-J., Hsu, J.-W., Huang, K.-L., Tu, P.-C., and Bai, Y.-M. (2022). Neurofilament light chain is a novel biomarker for major depression and related executive dysfunction. Int. J. Neuropsychopharmacol. 25, 99–105.

126. Brodaty, H., and Donkin, M. (2009). Family caregivers of people with dementia. Dialogues Clin Neurosci 11, 217–228.

127. Lindeza, P., Rodrigues, M., Costa, J., Guerreiro, M., and Rosa, M.M. (2020). Impact of dementia on informal care: a systematic review of family caregivers’ perceptions. BMJ Support. Palliat. Care.

128. Victor, C.R., Rippon, I., Quinn, C., Nelis, S.M., Martyr, A., Hart, N., Lamont, R., and Clare, L. (2021). The prevalence and predictors of loneliness in caregivers of people with dementia: findings from the IDEAL programme. Aging Ment. Health 25, 1232–1238.

129. Huang, S.-S. (2022). Depression among caregivers of patients with dementia: Associative factors and management approaches. World J. Psychiatry 12, 59–76.

130. Chesney, E., Goodwin, G.M., and Fazel, S. (2014). Risks of all-cause and suicide mortality in mental disorders: a meta-review. World Psychiatry 13, 153–160.

131. Fry, A., Littlejohns, T.J., Sudlow, C., Doherty, N., Adamska, L., Sprosen, T., Collins, R., and Allen, N.E. (2017). Comparison of Sociodemographic and Health-Related Characteristics of UK Biobank Participants With Those of the General Population. Am. J. Epidemiol. 186, 1026–1034.

132. Brommelhoff, J.A., Gatz, M., Johansson, B., McArdle, J.J., Fratiglioni, L., and Pedersen, N.L. (2009). Depression as a risk factor or prodromal feature for dementia? Findings in a population-based sample of Swedish twins. Psychol. Aging 24, 373–384.

133. Creese, B., and Ismail, Z. (2022). Mild behavioral impairment: measurement and clinical correlates of a novel marker of preclinical Alzheimer’s disease. Alzheimers Res. Ther. 14, 2.

134. DeMichele-Sweet, M.A.A., Klei, L., Creese, B., Harwood, J.C., Weamer, E.A., McClain, L., Sims, R., Hernandez, I., Moreno-Grau, S., Tárraga, L., et al. (2021). Genome-wide association identifies the first risk loci for psychosis in Alzheimer disease. Mol. Psychiatry 26, 5797–5811.

