## Supplementary Materials for "Investigating the genetic relationship between depression symptoms and Alzheimer’s Disease in clinically diagnosed and proxy cases"

**Supplementary Material**

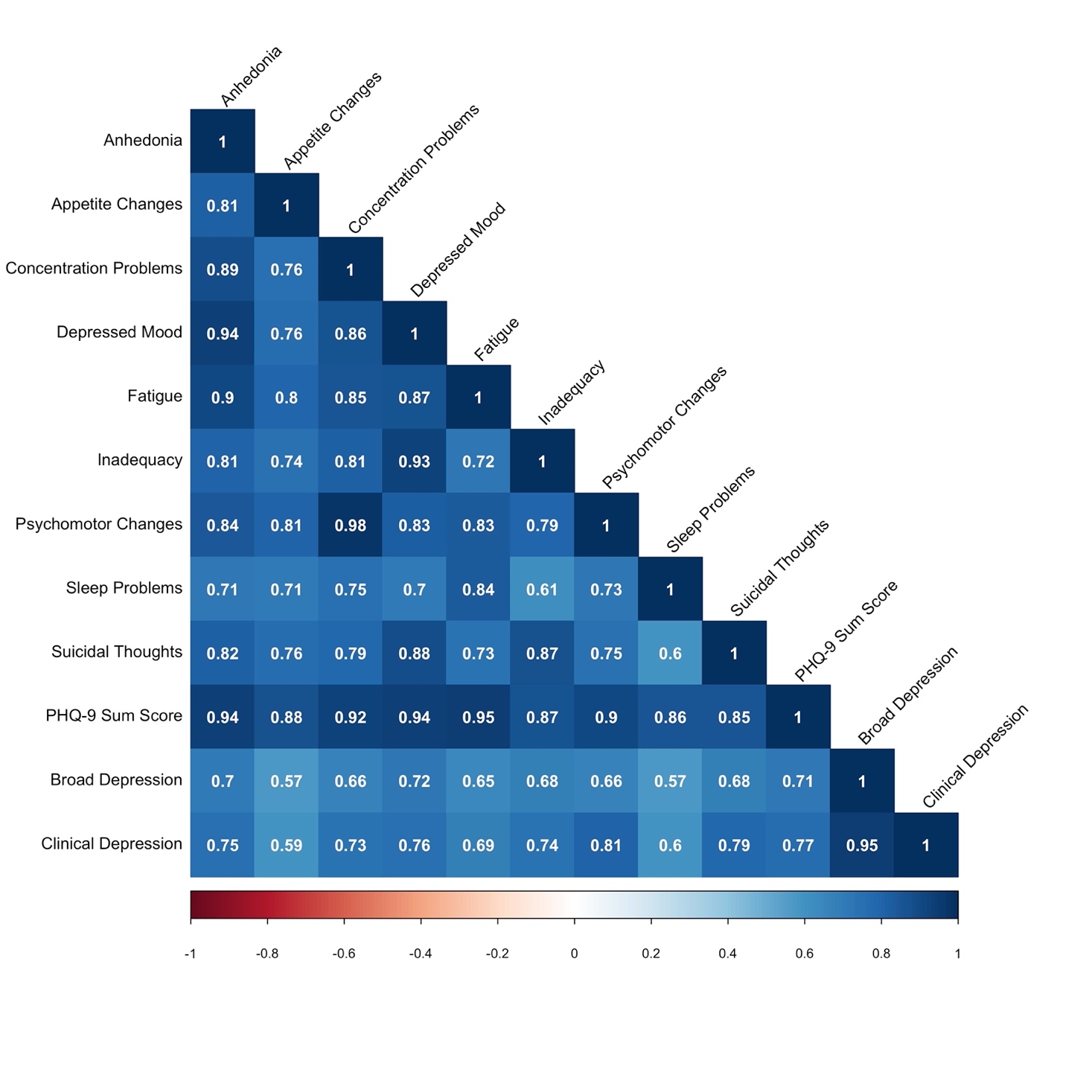
**Supplementary Material 1: Heatmap of depression phenotype genetic correlations**

Supplementary Material 1: Heatmap of LDSC calculated genetic correlation between the ten PHQ-9 depression phenotypes, clinical and broad depression

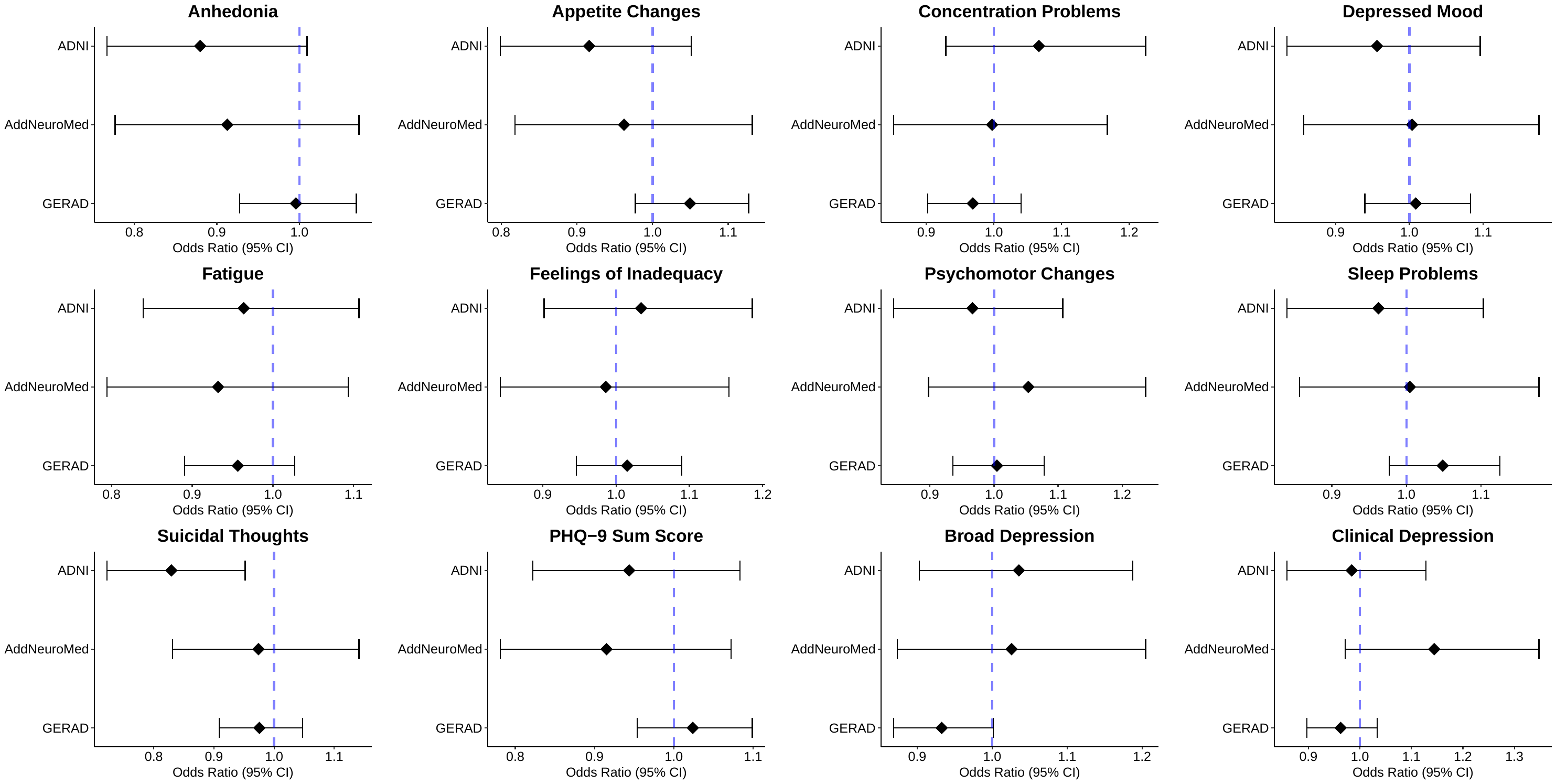
**Supplementary Material 2: Forest plot of polygenic risk score results in the three clinical Alzheimer’s cohorts**

Supplementary Material 2: Forest plots of polygenic risk score results for each depression phenotype in each of the three clinical Alzheimer’s Disease cohorts (including the *APOE* region)

**Supplementary Material 3: Genetic and Environmental Risk for Alzheimer’s disease Consortium (GERAD1) Collaborators**

**Individuals:**

Denise Harold^1^, Rebecca Sims^1^, Amy Gerrish^1^, Jade Chapman^1^, Valentina Escott- Price^1^, Nandini Badarinarayan^1^, Richard Abraham^1^, Paul Hollingworth^1^, Marian Hamshere^1^, Jaspreet Singh Pahwa^1^, Kimberley Dowzell^1^, Amy Williams^1^, Nicola Jones^1^, Charlene Thomas^1^, Alexandra Stretton^1^, Angharad Morgan^1^, Kate Williams^1^, Sarah Taylor^1^, Simon Lovestone^2^, John Powell^3^, Petroula Proitsi^3^, Michelle K Lupton^3^, Carol Brayne^4^, David C. Rubinsztein^5^, Michael Gill^6^, Brian Lawlor^6^, Aoibhinn Lynch^6^, Kevin Morgan^7^, Kristelle Brown^7^, Peter Passmore^8^, David Craig^8^, Bernadette McGuinness^8^, Janet A Johnston^8^, Stephen Todd^8^, Clive Holmes^9^, David Mann^10^, A. David Smith^11^, Seth Love^12^, Patrick G. Kehoe^12^, John Hardy^13^, Rita Guerreiro^14,15^, Andrew Singleton^14^, Simon Mead^16^, Nick Fox^17^, Martin Rossor^17^, John Collinge^16^, Wolfgang Maier^18^, Frank Jessen^18^, Reiner Heun^18^, Britta Schürmann^18,19^, Alfredo Ramirez^18^, Tim Becker^20^, Christine Herold^20^, André Lacour^20^, Dmitriy Drichel^20^, Hendrik van den Bussche^21^, Isabella Heuser^22^, Johannes Kornhuber^23^, Jens Wiltfang^24^, Martin Dichgans^25,26^, Lutz Frölich^27^, Harald Hampel^28^, Michael Hüll^29^, Dan Rujescu^30^, Alison Goate^31^, John S.K. Kauwe^32^, Carlos Cruchaga^33^, Petra Nowotny^33^, John C. Morris^33^, Kevin Mayo^33^, Gill Livingston^34^, Nicholas J. Bass^34^, Hugh Gurling^34^, Andrew McQuillin^34^, Rhian Gwilliam^35^, Panagiotis Deloukas^35^, Markus M. Nöthen^20^, Peter Holmans^1^, Michael O’Donovan^1^, Michael J.Owen^1^, Julie Williams^1^.

**Affiliations:**

^1^Medical Research Council (MRC) Centre for Neuropsychiatric Genetics and Genomics, Neurosciences and Mental Health Research Institute, Department of Psychological Medicine and Neurology, School of Medicine, Cardiff University, Cardiff, UK. ^2^Department of Psychiatry, Medical Sciences Division, University of Oxford, Oxford, UK. ^3^Kings College London, Institute of Psychiatry, Department of Neuroscience, De Crespigny Park, Denmark Hill, London, UK. ^4^Institute of Public Health, University of Cambridge, Cambridge, UK. ^5^Cambridge Institute for Medical Research, University of Cambridge, Cambridge, UK. ^6^Mercers Institute for Research on Aging, St. James Hospital and Trinity College, Dublin, Ireland. ^7^Institute of Genetics, Queens Medical Centre, University of Nottingham, UK. ^8^Ageing Group, Centre for Public Health, School of Medicine, Dentistry and Biomedical Sciences, Queens University Belfast, UK. ^9^Division of Clinical Neurosciences, School of Medicine, University of Southampton, Southampton, UK. ^10^Clinical Neuroscience Research Group, Greater Manchester Neurosciences Centre, University of Manchester, Salford, UK. ^11^Oxford Project to Investigate Memory and Ageing (OPTIMA), University of Oxford, Department of Pharmacology, Mansfield Road, Oxford, UK. ^12^University of Bristol Institute of Clinical Neurosciences, School of Clinical Sciences, Frenchay Hospital, Bristol, UK. ^13^Department of Molecular Neuroscience and Reta Lilla Weston Laboratories, Institute of Neurology, UCL, London, UK. ^14^Laboratory of Neurogenetics, National Institute on Aging, National Institutes of Health, Bethesda, Maryland, United States of America. ^15^Department of Molecular Neuroscience, Institute of Neurology, University College London, Queen Square, London WC1N 3BG, UK. ^16^MRC Prion Unit, Department of Neurodegenerative Disease, UCL Institute of Neurology, London, UK. ^17^Dementia Research Centre, Department of Neurodegenerative Diseases, University College London, Institute of Neurology, London, UK. ^18^Department of Psychiatry, University of Bonn, Sigmund-

Freud-Straβe 25, 53105 Bonn, Germany. ^19^Institute for Molecular Psychiatry, University of Bonn, Bonn, Germany. ^20^Department of Genomics, Life & Brain Center, University of Bonn, Bonn, Germany. ^21^Institute of Primary Medical Care, University Medical Center Hamburg-Eppendorf, Germany. ^22^Department of Psychiatry, Charité Berlin, Germany. ^23^Department of Psychiatry, Friedrich-Alexander-University Erlangen-Nürnberg, Germany. ^24^Department of Psychiatry and Psychotherapy, University Medical Center (UMG), Georg-August-University, Göttingen, Germany. ^25^Institute for Stroke and Dementia Research, Klinikum der Universität München, Marchioninistr. 15, 81377, Munich, Germany. ^26^Department of Neurology, Klinikum der Universität München, Marchioninistr. 15, 81377, Munich, Germany. ^27^Central Institute of Mental Health, Medical Faculty Mannheim, University of Heidelberg, Germany. ^28^Institute for Memory and Alzheimer’s Disease & INSERM, Sorbonne Universities, Pierre and Marie Curie University, Paris, France; Institute for Brain and Spinal Cord Disorders (ICM), Department of Neurology, Hospital of Pitié-Salpétrière, Paris, France. ^29^Centre for Geriatric Medicine and Section of Gerontopsychiatry and Neuropsychology, Medical School, University of Freiburg, Germany. ^30^Department of Psychiatry, University of Halle, Halle, Germany. ^31^Neuroscience Department, Icahn School of Medicine at Mount Sinai, New York, US. ^32^Department of Biology, Brigham Young University, Provo, UT, 84602, USA. ^33^Departments of Psychiatry, Neurology and Genetics, Washington University School of Medicine, St Louis, MO 63110, US. ^34^Department of Mental Health Sciences, University College London, UK. ^35^The Wellcome Trust Sanger Institute, Wellcome Trust Genome Campus, Hinxton, Cambridge, UK.
